## Supplemental Figure S1 for "Ventilation during COVID-19 in a school for students with intellectual and developmental disabilities (IDD)"

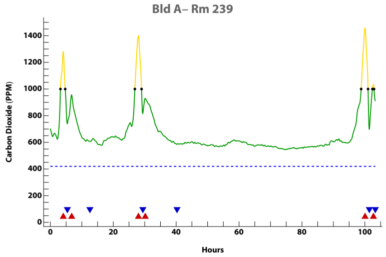


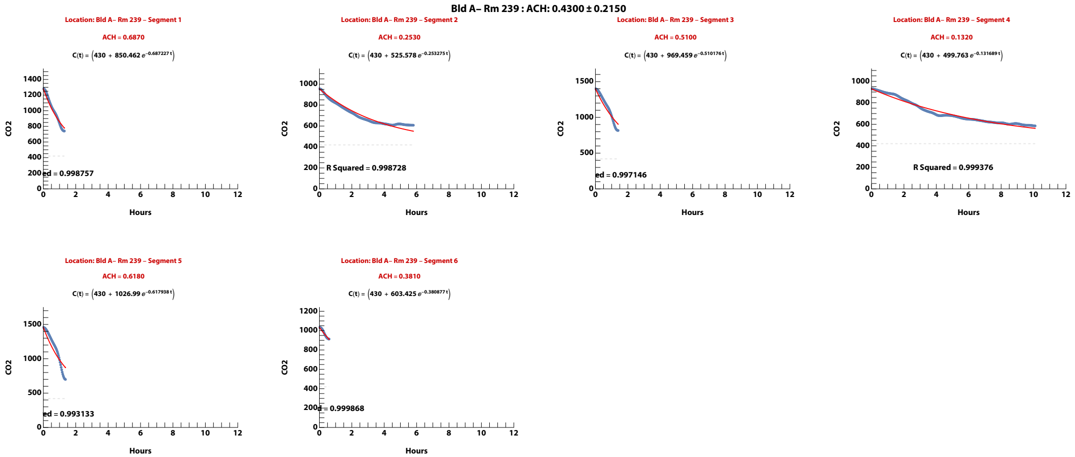


**Zand, et al. (2023). Ventilation during COVID–19 in a school for students with intellectual and developmental disabilities (IDD) [Submitted]**

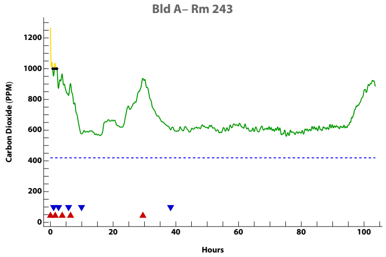


Bld A– Rm 243 : ACH: 0.2820 ± 0.1850

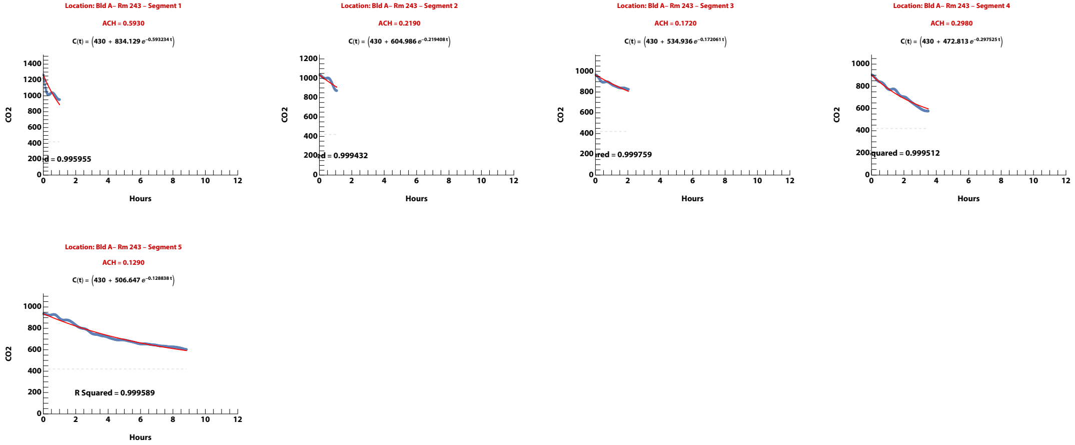


**Zand, et al. (2023). Ventilation during COVID–19 in a school for students with intellectual and developmental disabilities (IDD) [Submitted]**

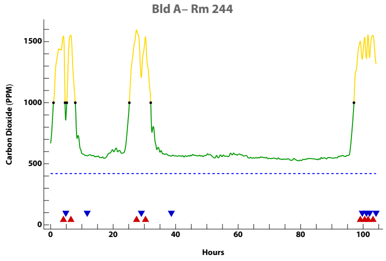


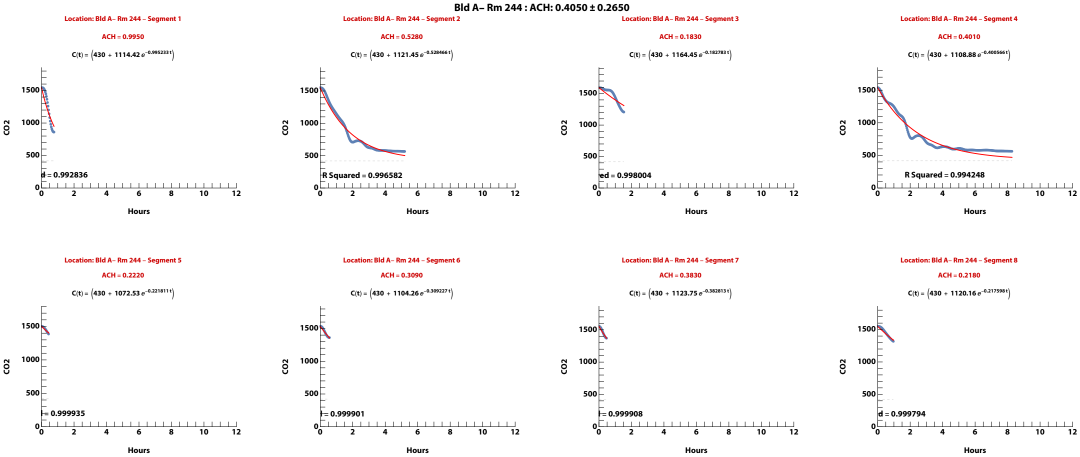


**Zand, et al. (2023). Ventilation during COVID–19 in a school for students with intellectual and developmental disabilities (IDD) [Submitted]**

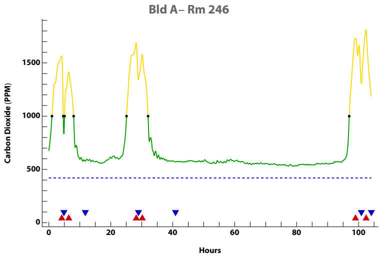


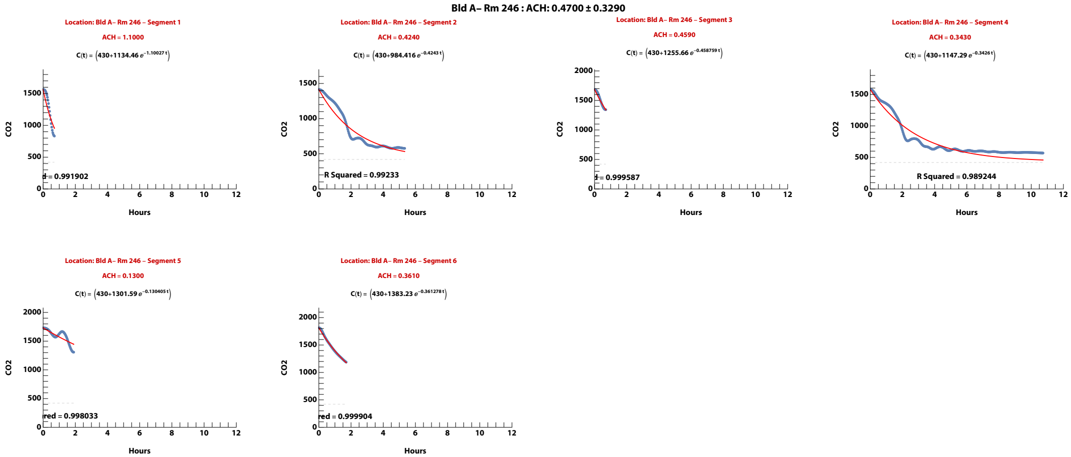


**Zand, et al. (2023). Ventilation during COVID–19 in a school for students with intellectual and developmental disabilities (IDD) [Submitted]**

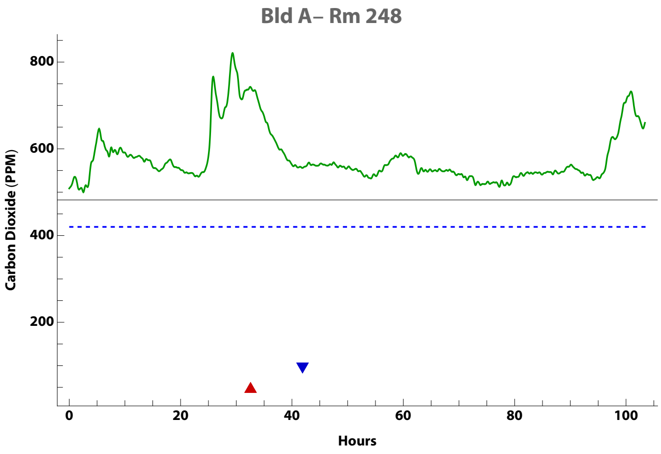


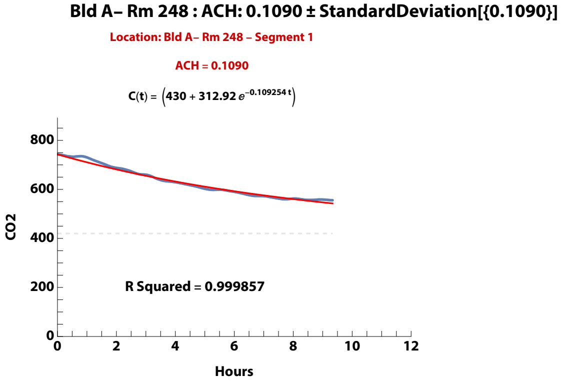


**Zand, et al. (2023). Ventilation during COVID–19 in a school for students with intellectual and developmental disabilities (IDD) [Submitted]**

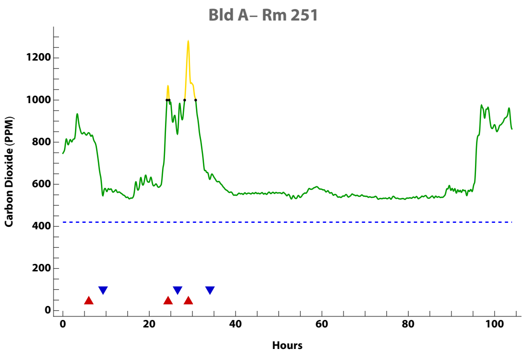


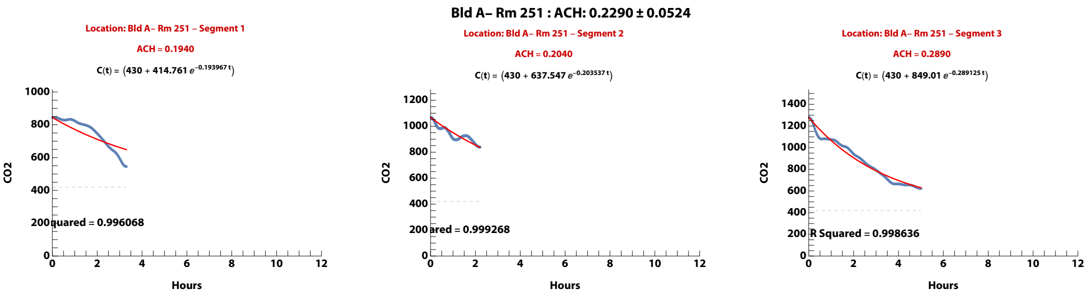


**Zand, et al. (2023). Ventilation during COVID–19 in a school for students with intellectual and developmental disabilities (IDD) [Submitted]**

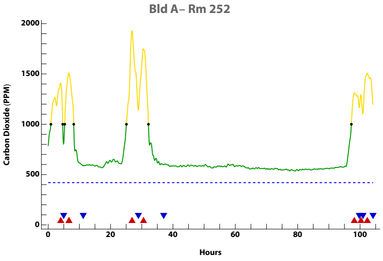


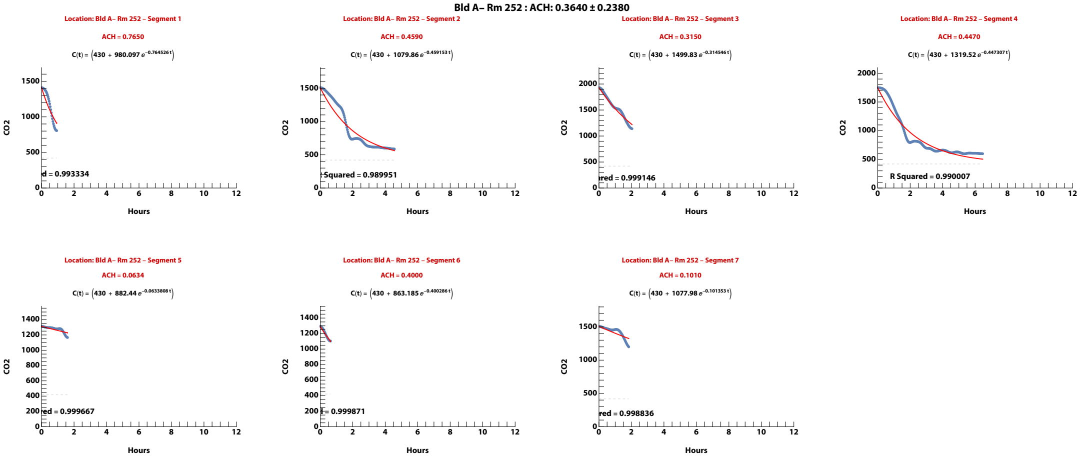


**Zand, et al. (2023). Ventilation during COVID–19 in a school for students with intellectual and developmental disabilities (IDD) [Submitted]**

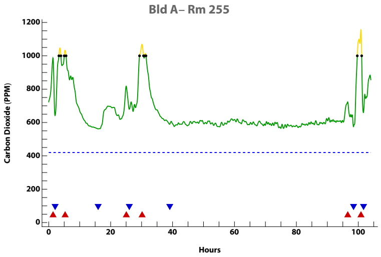


Bld A– Rm 255 : ACH: 0.6450 ± 0.5710

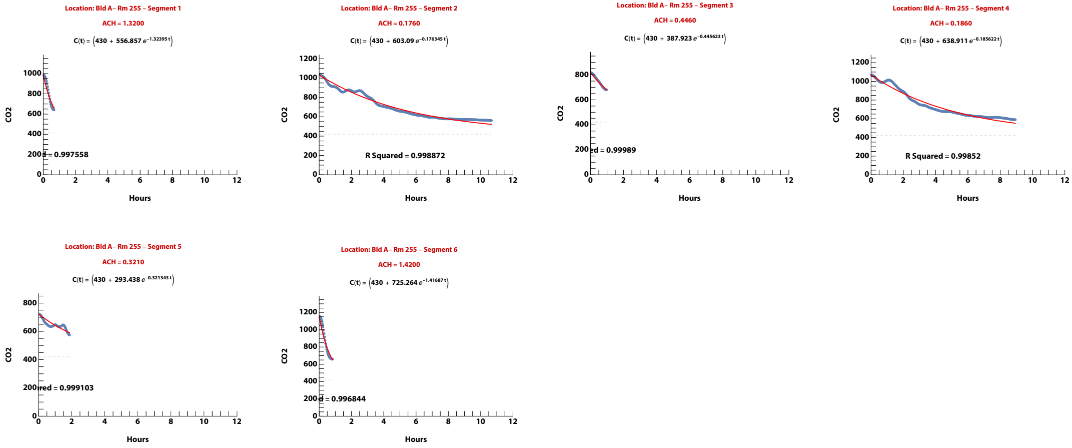


**Zand, et al. (2023). Ventilation during COVID–19 in a school for students with intellectual and developmental disabilities (IDD) [Submitted]**

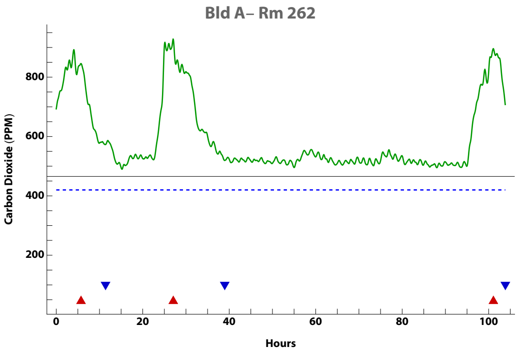


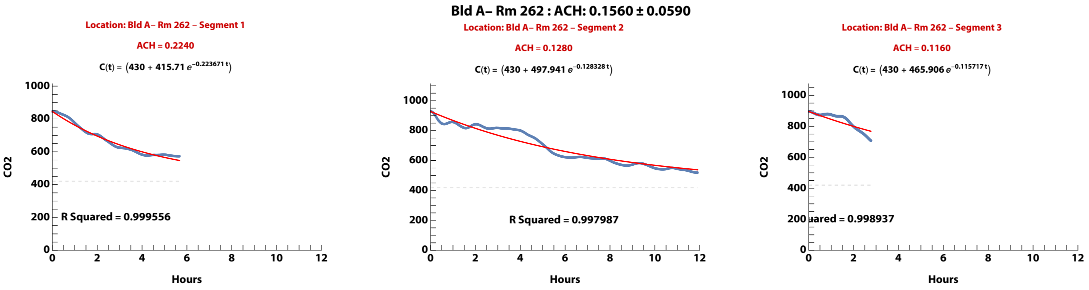


**Zand, et al. (2023). Ventilation during COVID–19 in a school for students with intellectual and developmental disabilities (IDD) [Submitted]**

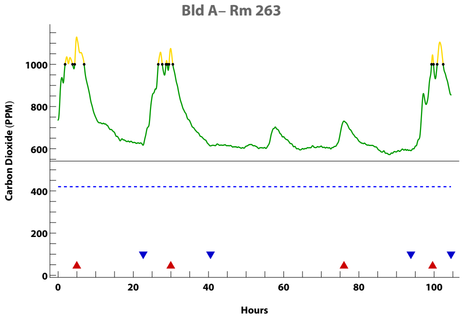


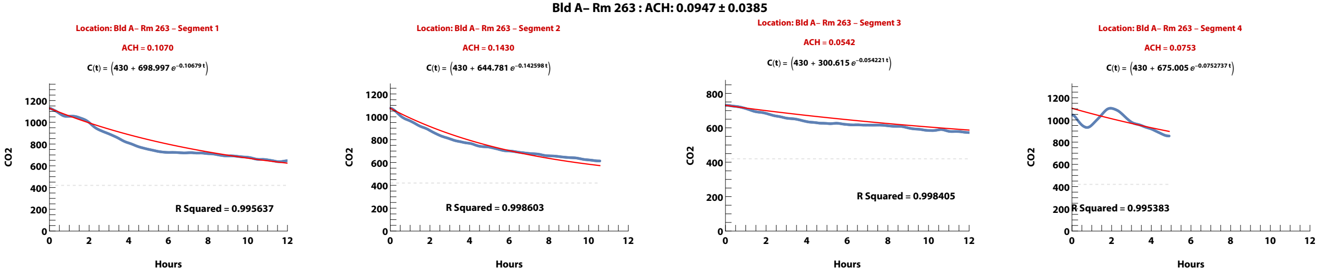


**Zand, et al. (2023). Ventilation during COVID–19 in a school for students with intellectual and developmental disabilities (IDD) [Submitted]**

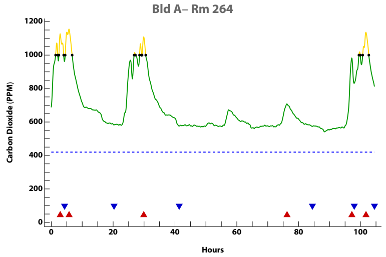


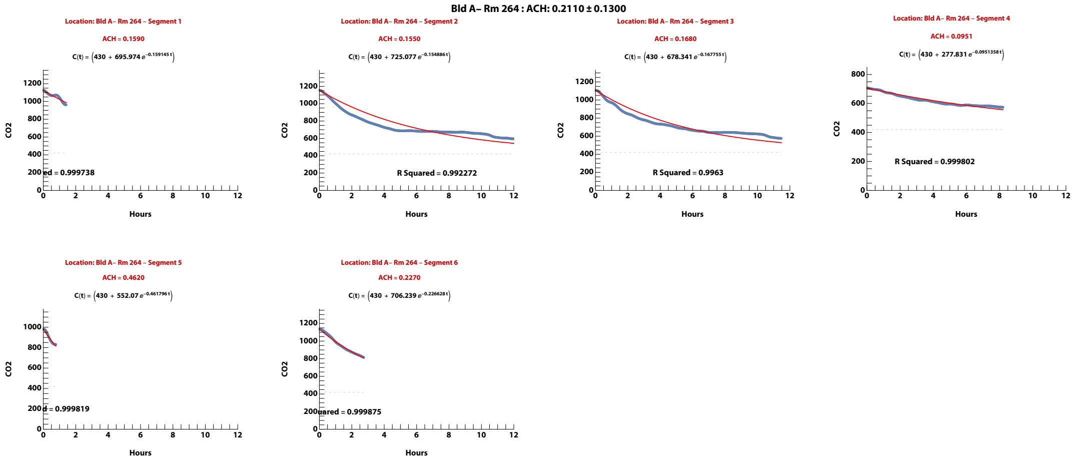


**Zand, et al. (2023). Ventilation during COVID–19 in a school for students with intellectual and developmental disabilities (IDD) [Submitted]**

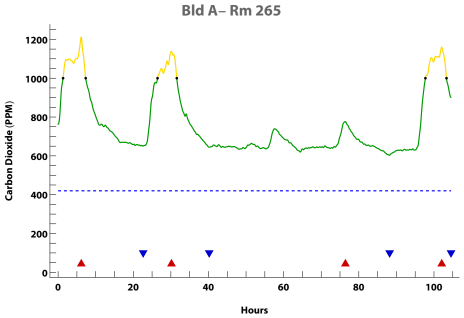


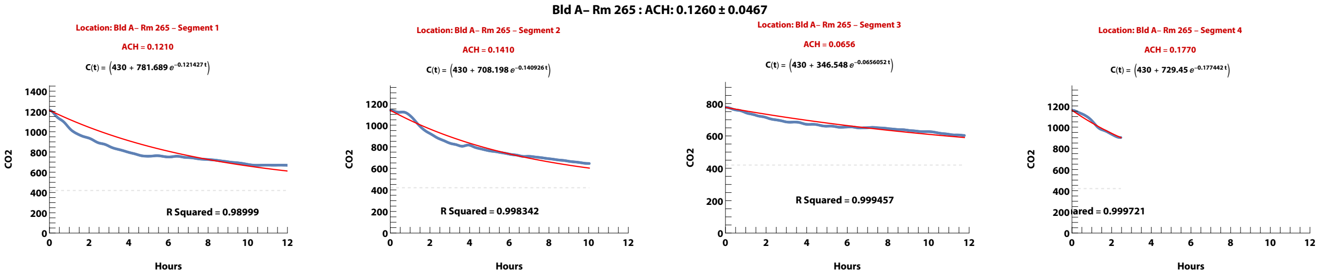


**Zand, et al. (2023). Ventilation during COVID–19 in a school for students with intellectual and developmental disabilities (IDD) [Submitted]**

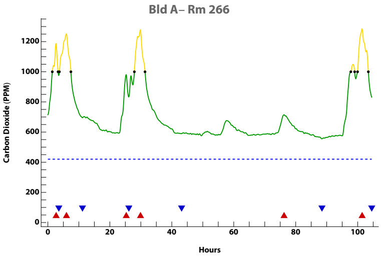


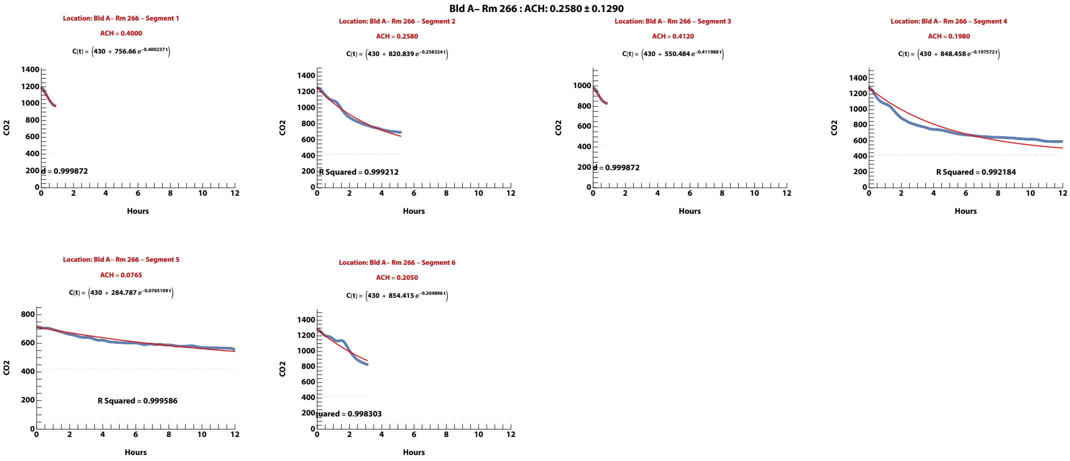


**Zand, et al. (2023). Ventilation during COVID–19 in a school for students with intellectual and developmental disabilities (IDD) [Submitted]**

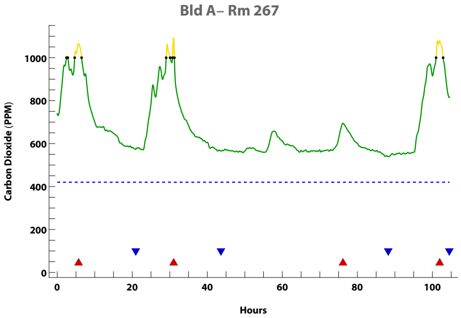


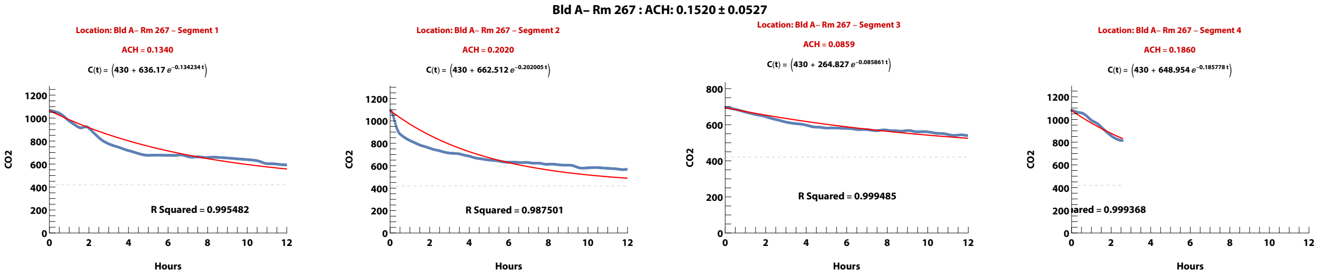


**Zand, et al. (2023). Ventilation during COVID–19 in a school for students with intellectual and developmental disabilities (IDD) [Submitted]**

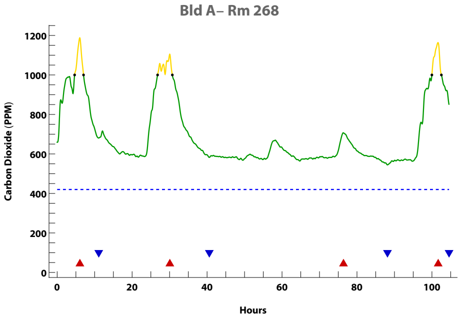


**Bld A– Rm 268 : ACH: 0.1730 ± 0.0683**

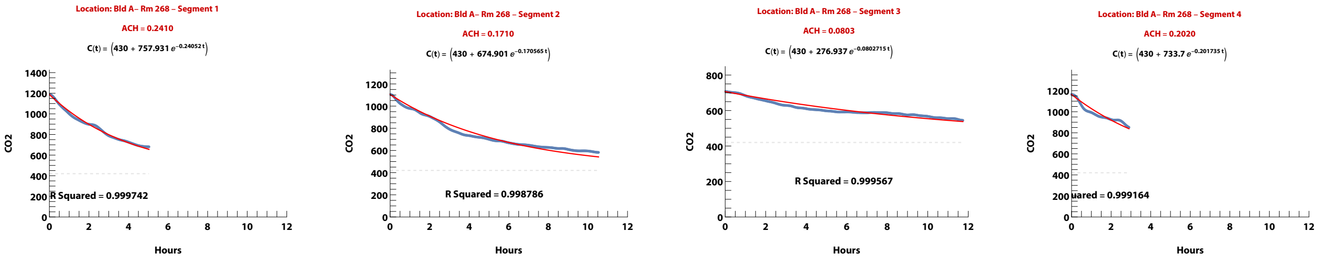


**Zand, et al. (2023). Ventilation during COVID–19 in a school for students with intellectual and developmental disabilities (IDD) [Submitted]**

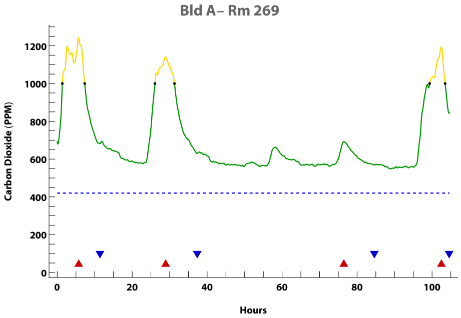


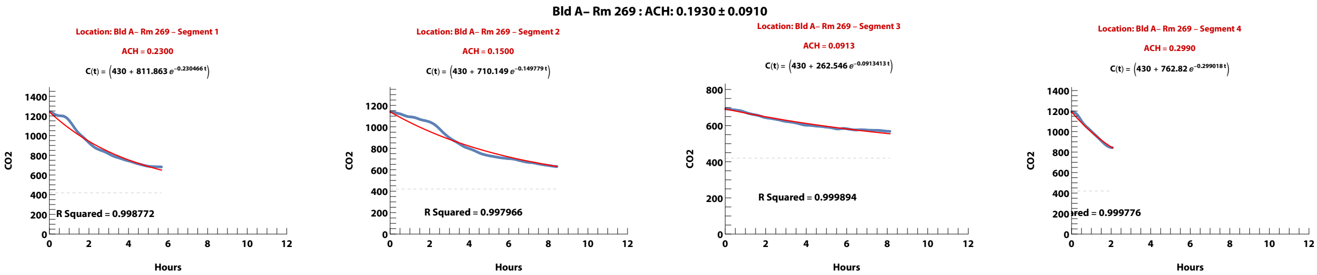


**Zand, et al. (2023). Ventilation during COVID–19 in a school for students with intellectual and developmental disabilities (IDD) [Submitted]**

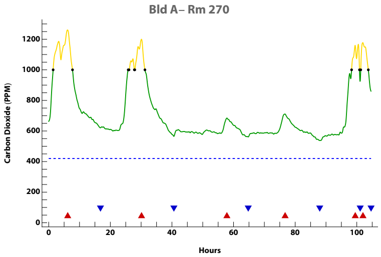


Bld A– Rm 270 : ACH: 0.1410 ± 0.0470

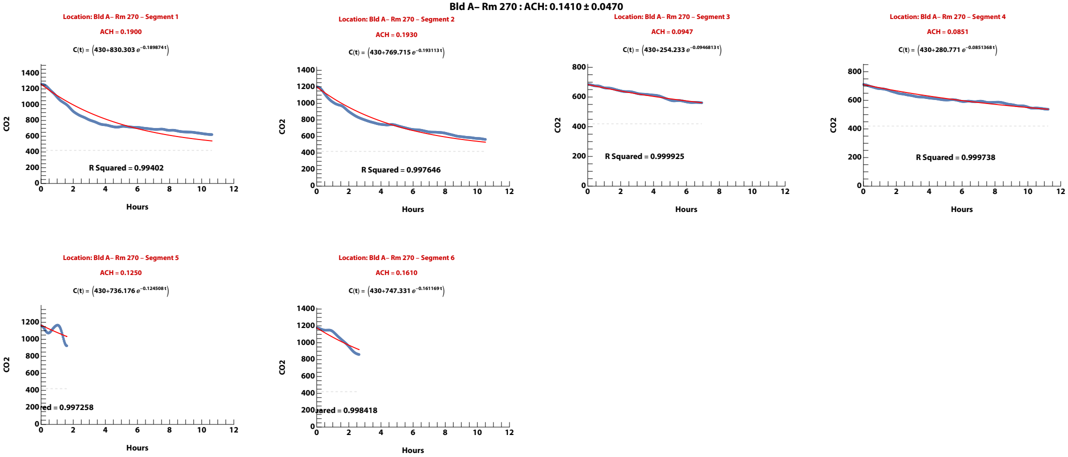


**Zand, et al. (2023). Ventilation during COVID–19 in a school for students with intellectual and developmental disabilities (IDD) [Submitted]**

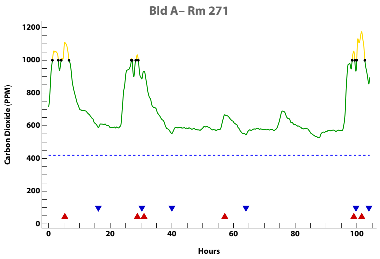


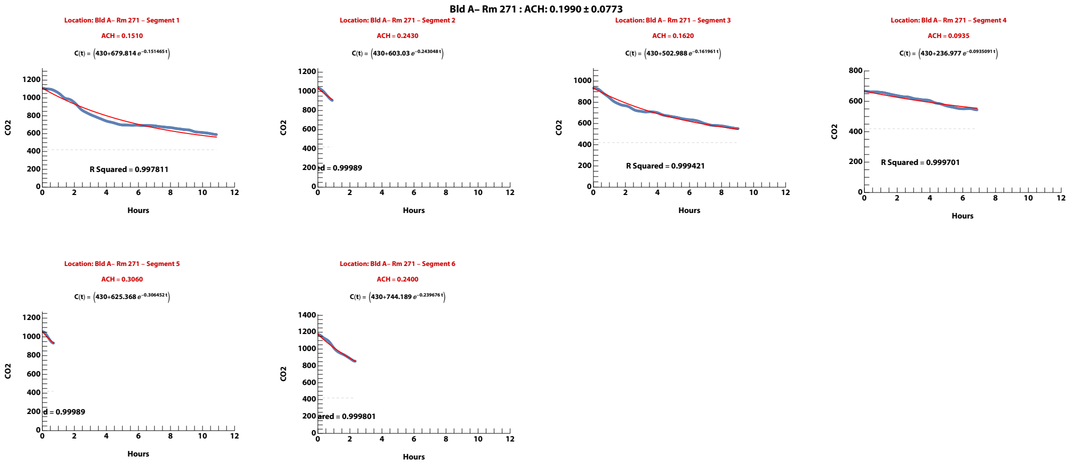


**Zand, et al. (2023). Ventilation during COVID–19 in a school for students with intellectual and developmental disabilities (IDD) [Submitted]**

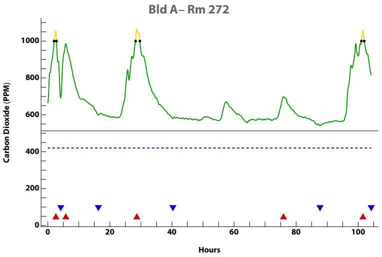


Bld A– Rm 272 : ACH: 0.1980 ± 0.1560

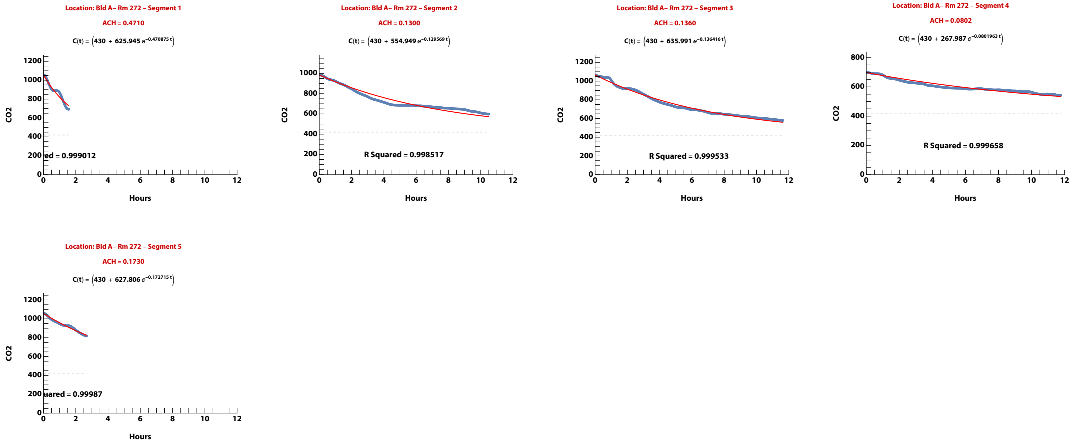


**Zand, et al. (2023). Ventilation during COVID–19 in a school for students with intellectual and developmental disabilities (IDD) [Submitted]**

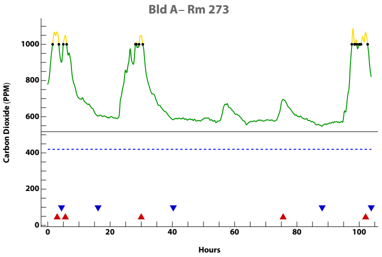


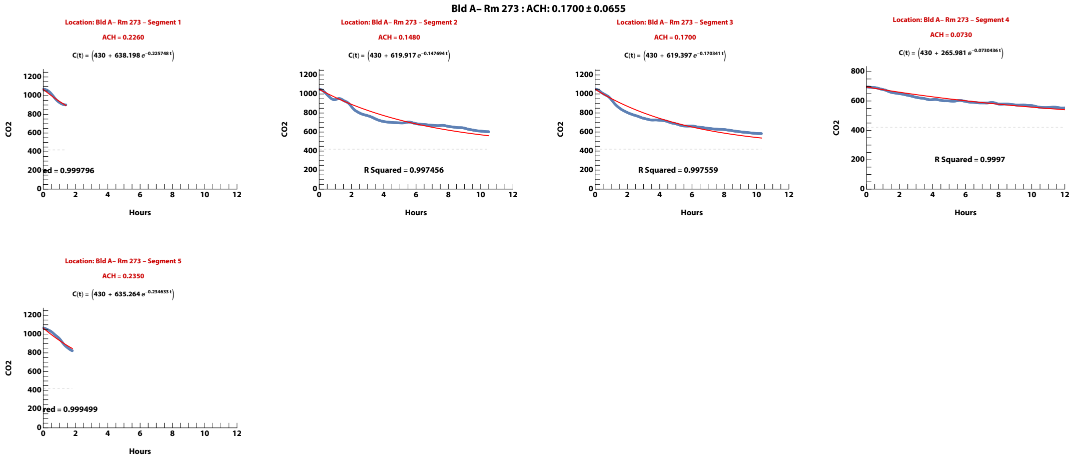


**Zand, et al. (2023). Ventilation during COVID–19 in a school for students with intellectual and developmental disabilities (IDD) [Submitted]**

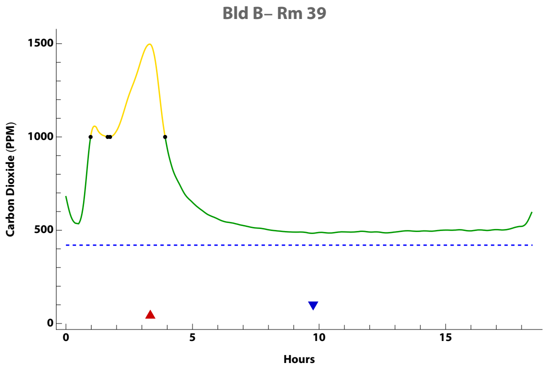


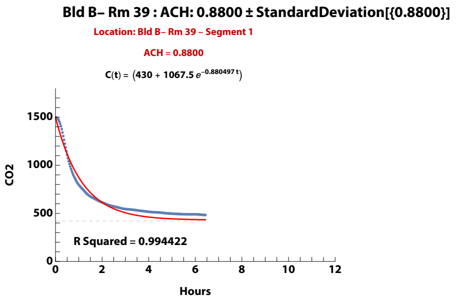


Zand, et al. (2023). Ventilation during COVID–19 in a school for students with intellectual and developmental disabilities (IDD) [Submitted]

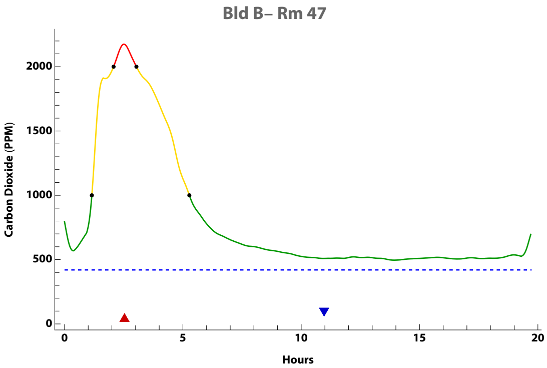


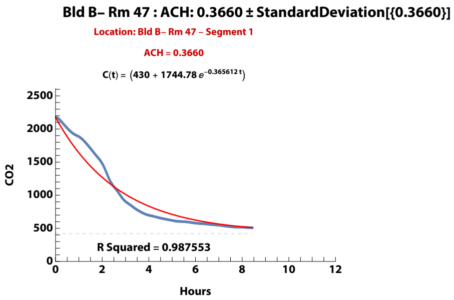


**Zand, et al. (2023). Ventilation during COVID–19 in a school for students with intellectual and developmental disabilities (IDD) [Submitted]**

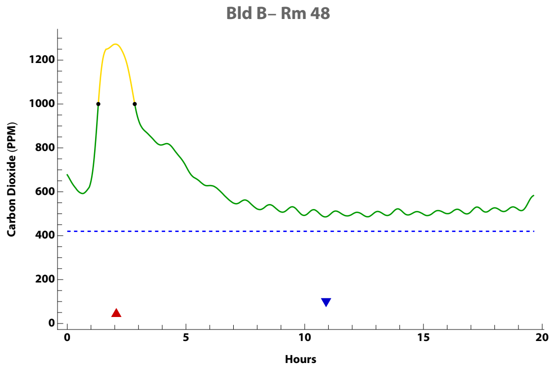


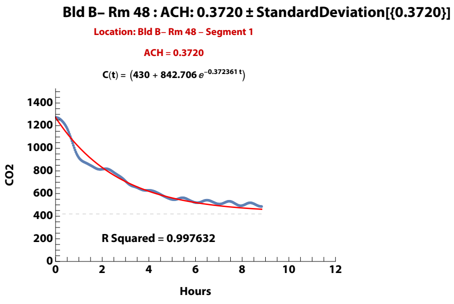


**Zand, et al. (2023). Ventilation during COVID–19 in a school for students with intellectual and developmental disabilities (IDD) [Submitted]**

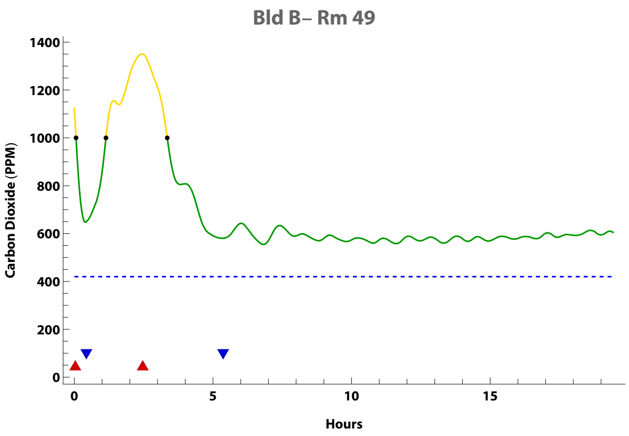


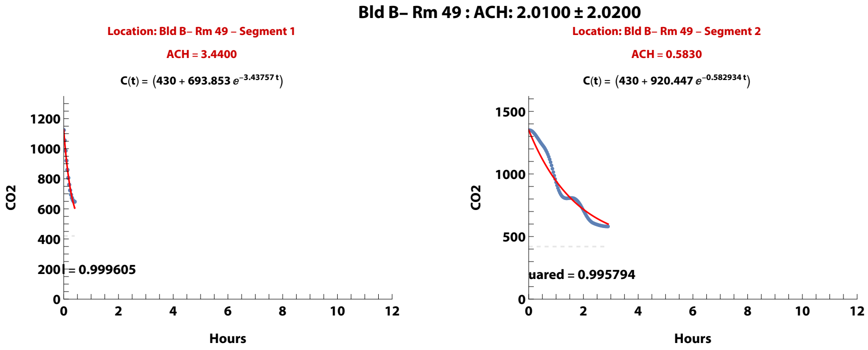


**Zand, et al. (2023). Ventilation during COVID–19 in a school for students with intellectual and developmental disabilities (IDD) [Submitted]**

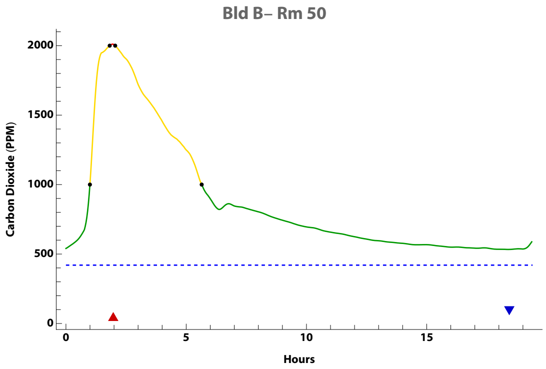


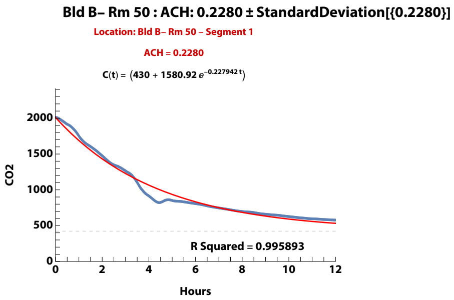


**Zand, et al. (2023). Ventilation during COVID–19 in a school for students with intellectual and developmental disabilities (IDD) [Submitted]**

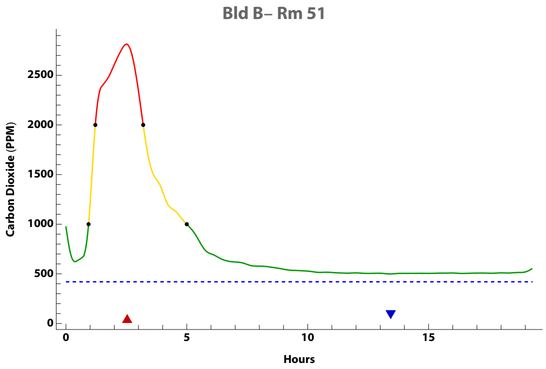


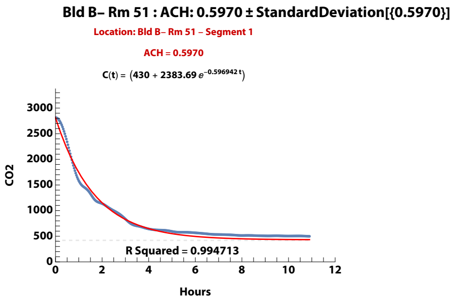


**Zand, et al. (2023). Ventilation during COVID–19 in a school for students with intellectual and developmental disabilities (IDD) [Submitted]**

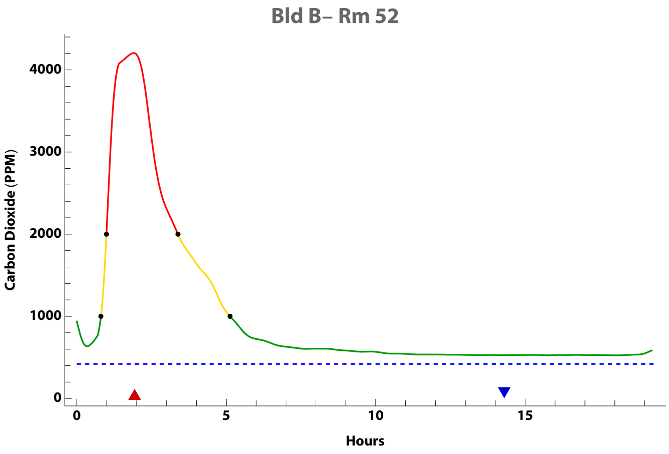


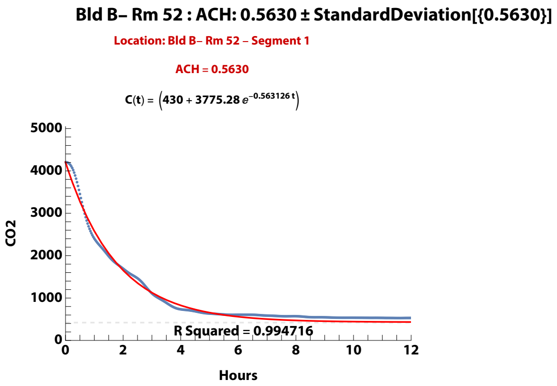


**Zand, et al. (2023). Ventilation during COVID–19 in a school for students with intellectual and developmental disabilities (IDD) [Submitted]**

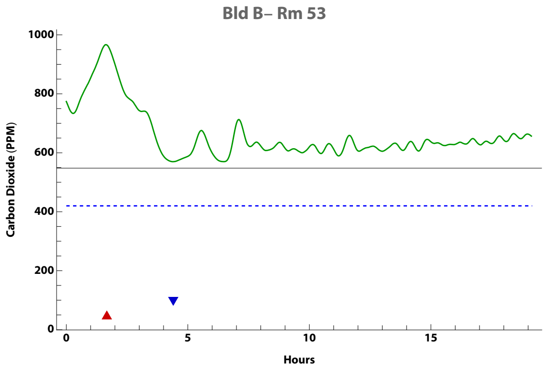


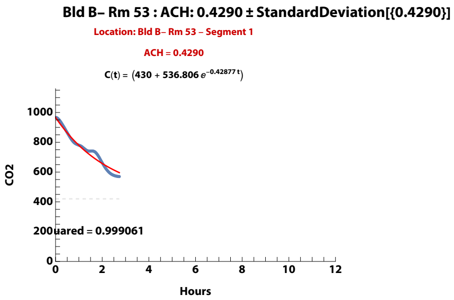


**Zand, et al. (2023). Ventilation during COVID–19 in a school for students with intellectual and developmental disabilities (IDD) [Submitted]**

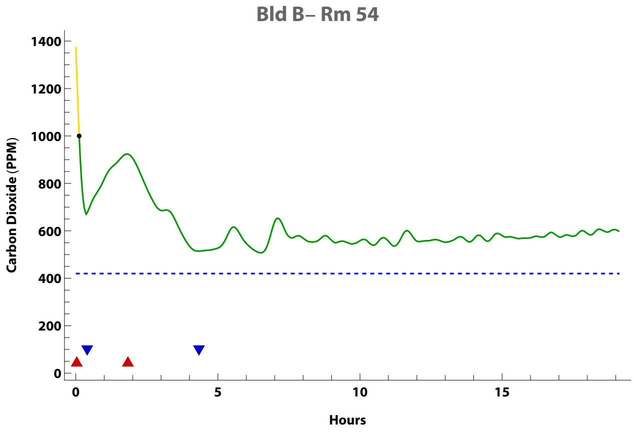


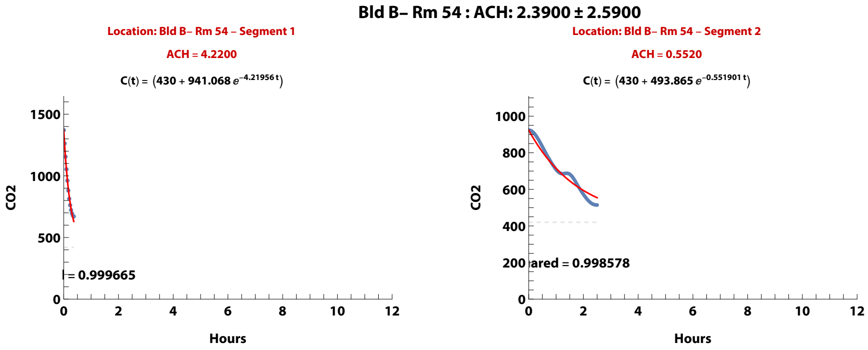


**Zand, et al. (2023). Ventilation during COVID–19 in a school for students with intellectual and developmental disabilities (IDD) [Submitted]**

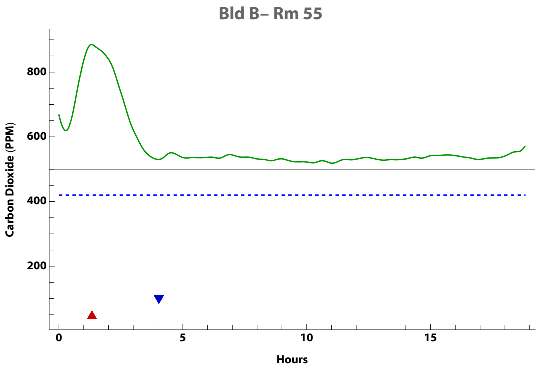


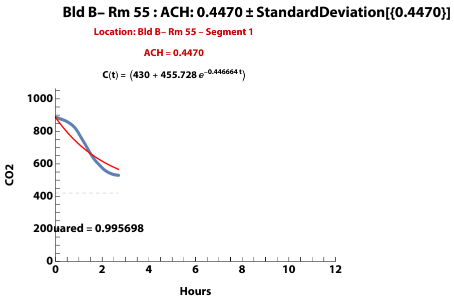


**Zand, et al. (2023). Ventilation during COVID–19 in a school for students with intellectual and developmental disabilities (IDD) [Submitted]**

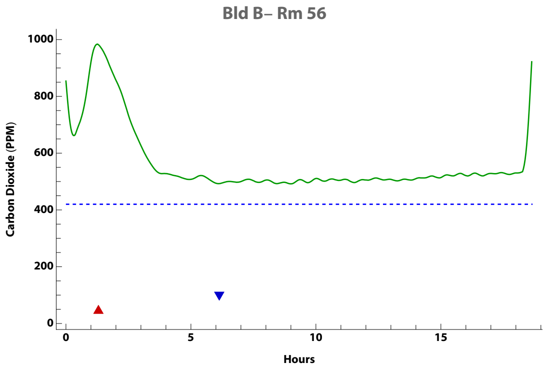


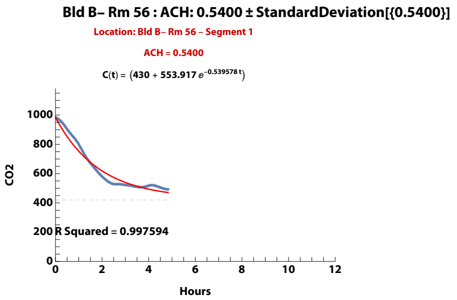


**Zand, et al. (2023). Ventilation during COVID–19 in a school for students with intellectual and developmental disabilities (IDD) [Submitted]**

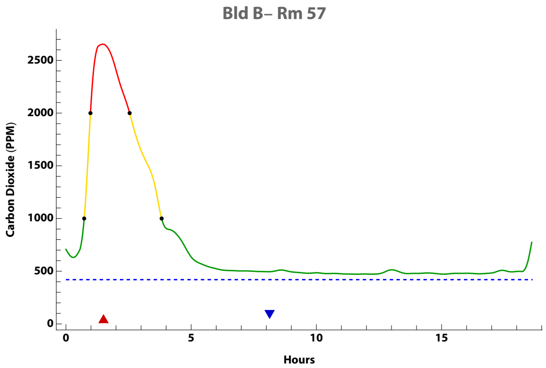


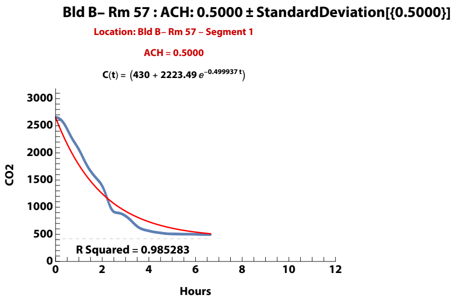


**Zand, et al. (2023). Ventilation during COVID–19 in a school for students with intellectual and developmental disabilities (IDD) [Submitted]**

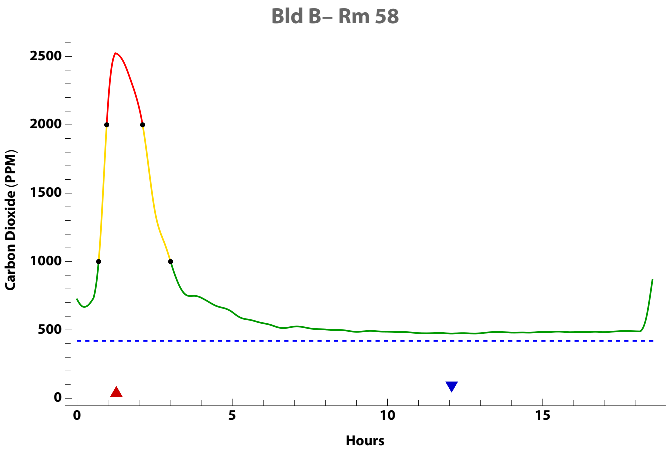


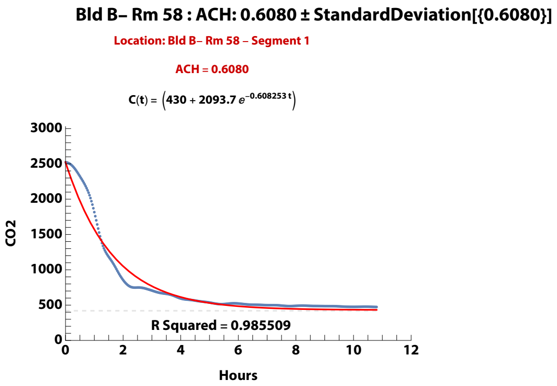


**Zand, et al. (2023). Ventilation during COVID–19 in a school for students with intellectual and developmental disabilities (IDD) [Submitted]**

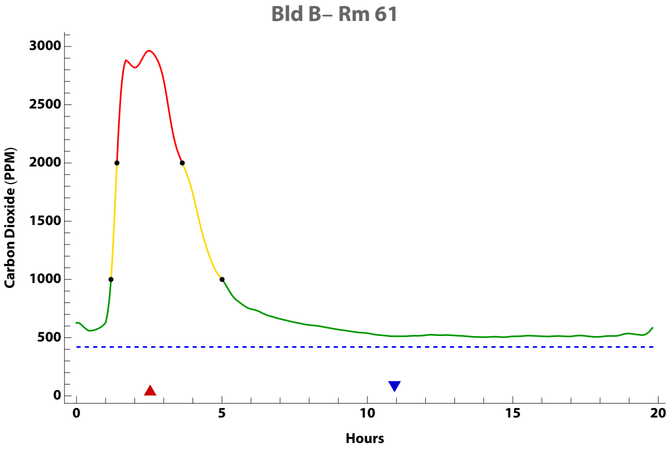


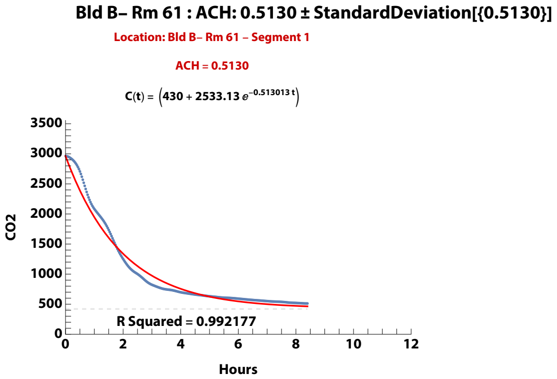


**Zand, et al. (2023). Ventilation during COVID–19 in a school for students with intellectual and developmental disabilities (IDD) [Submitted]**

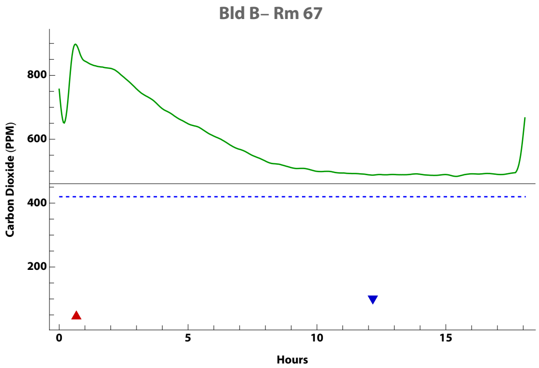


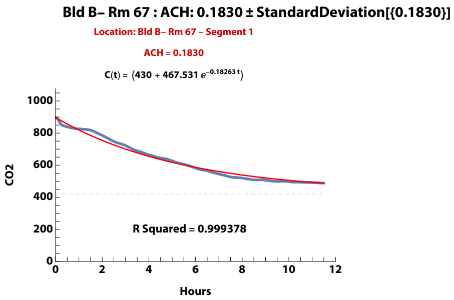


**Zand, et al. (2023). Ventilation during COVID–19 in a school for students with intellectual and developmental disabilities (IDD) [Submitted]**

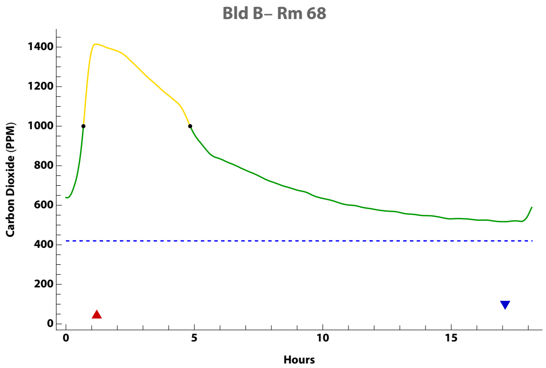


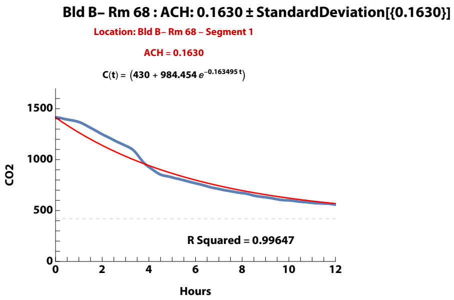


**Zand, et al. (2023). Ventilation during COVID–19 in a school for students with intellectual and developmental disabilities (IDD) [Submitted]**

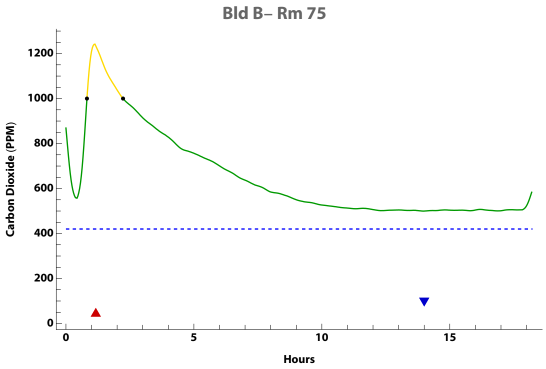


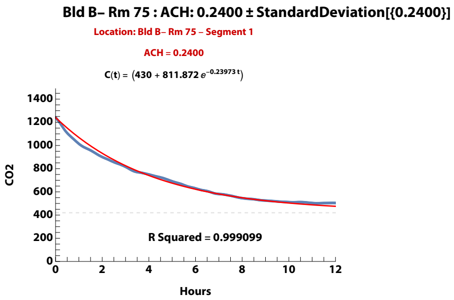


**Zand, et al. (2023). Ventilation during COVID–19 in a school for students with intellectual and developmental disabilities (IDD) [Submitted]**

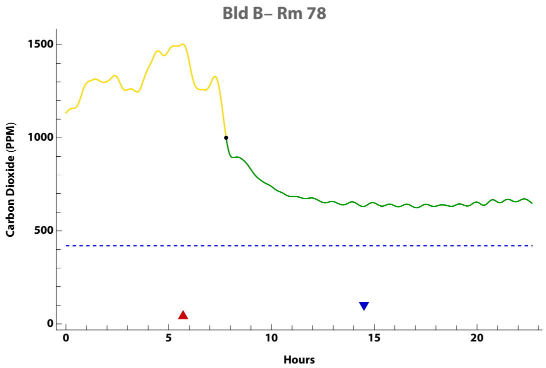


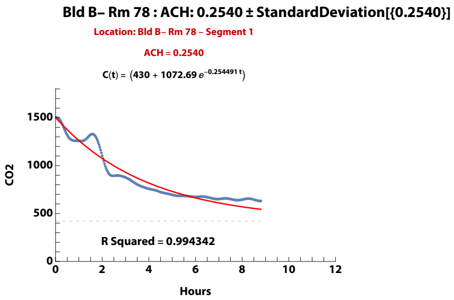


**Zand, et al. (2023). Ventilation during COVID–19 in a school for students with intellectual and developmental disabilities (IDD) [Submitted]**

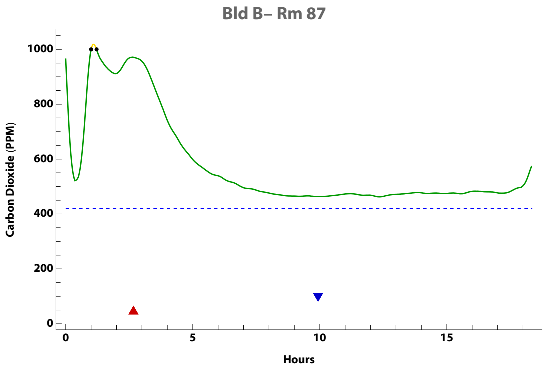


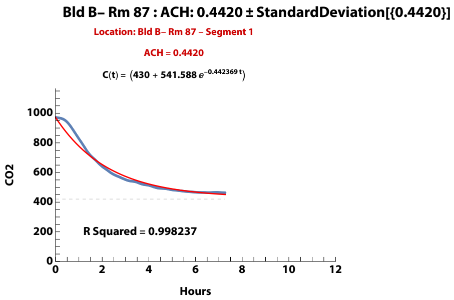


**Zand, et al. (2023). Ventilation during COVID–19 in a school for students with intellectual and developmental disabilities (IDD) [Submitted]**

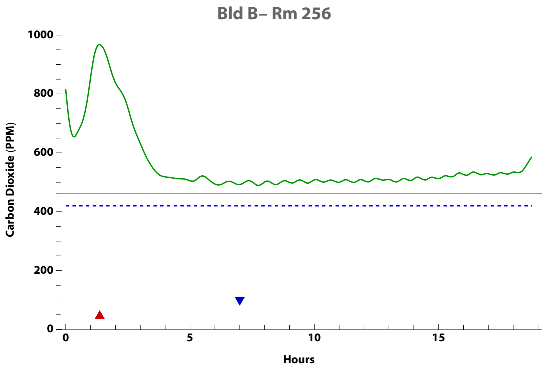


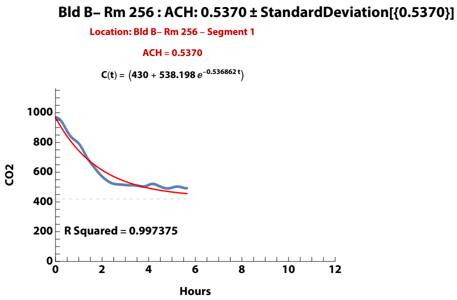


**Zand, et al. (2023). Ventilation during COVID–19 in a school for students with intellectual and developmental disabilities (IDD) [Submitted]**

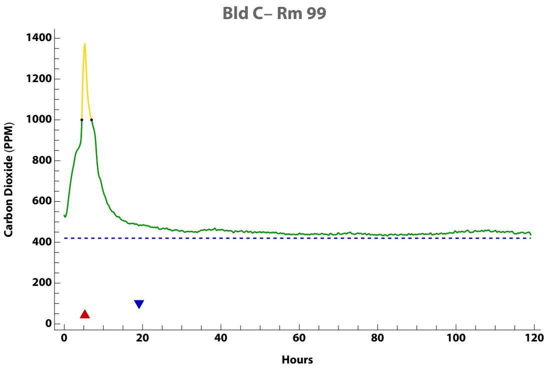


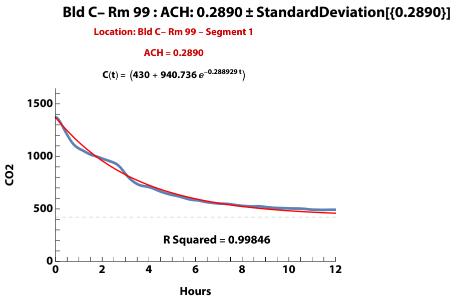


**Zand, et al. (2023). Ventilation during COVID–19 in a school for students with intellectual and developmental disabilities (IDD) [Submitted]**

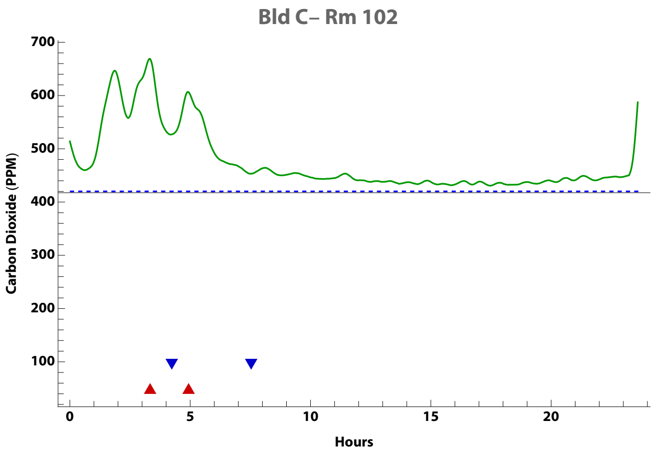


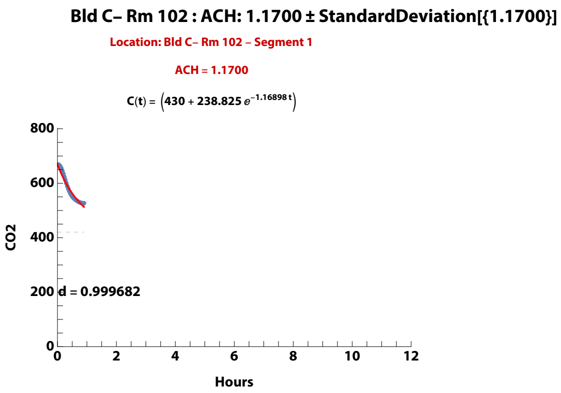


**Zand, et al. (2023). Ventilation during COVID–19 in a school for students with intellectual and developmental disabilities (IDD) [Submitted]**

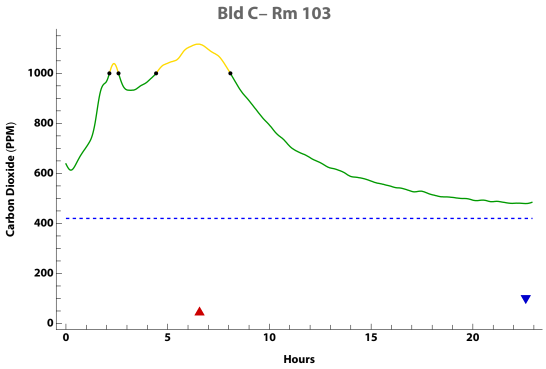


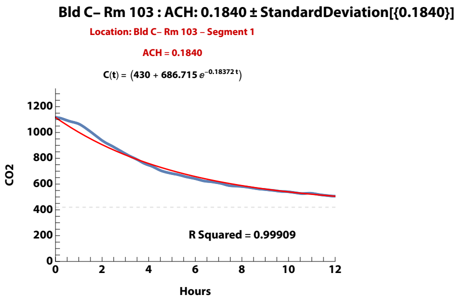


**Zand, et al. (2023). Ventilation during COVID–19 in a school for students with intellectual and developmental disabilities (IDD) [Submitted]**

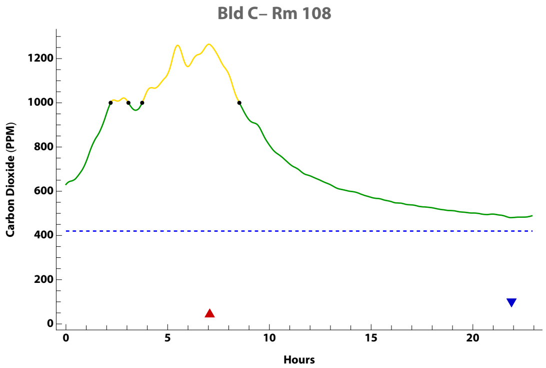


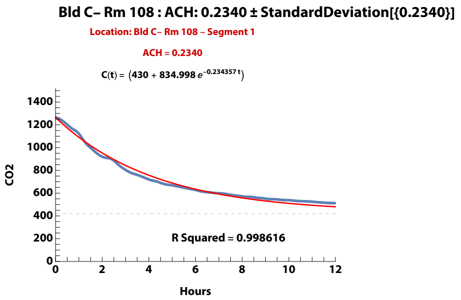


**Zand, et al. (2023). Ventilation during COVID–19 in a school for students with intellectual and developmental disabilities (IDD) [Submitted]**

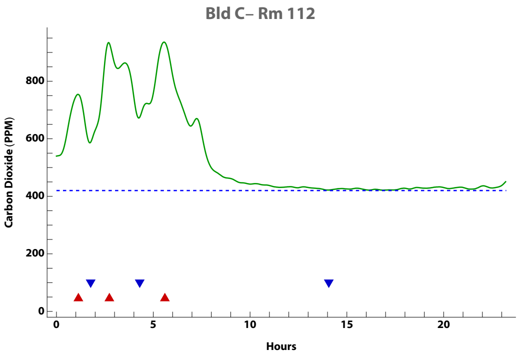


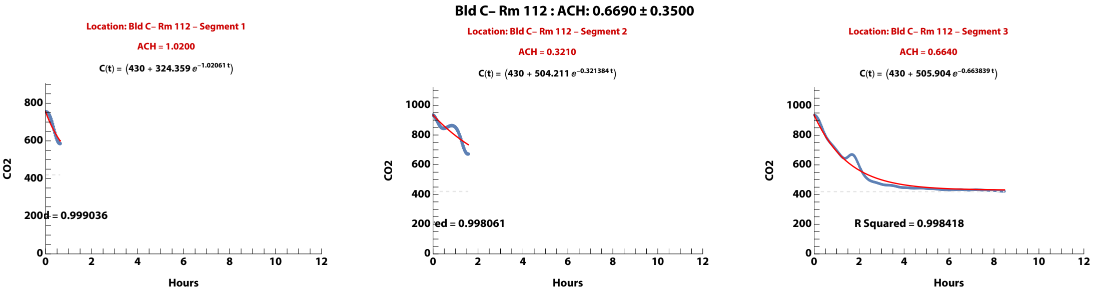


**Zand, et al. (2023). Ventilation during COVID–19 in a school for students with intellectual and developmental disabilities (IDD) [Submitted]**

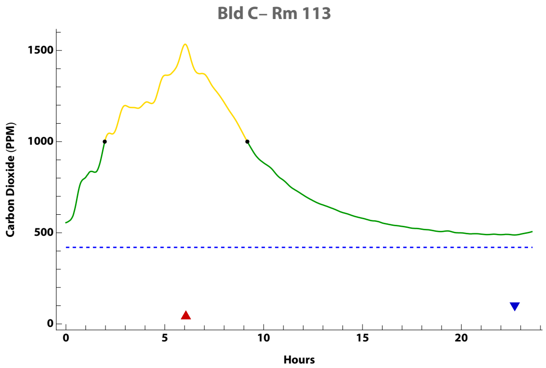


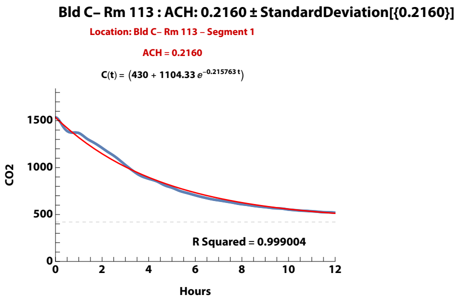


**Zand, et al. (2023). Ventilation during COVID–19 in a school for students with intellectual and developmental disabilities (IDD) [Submitted]**

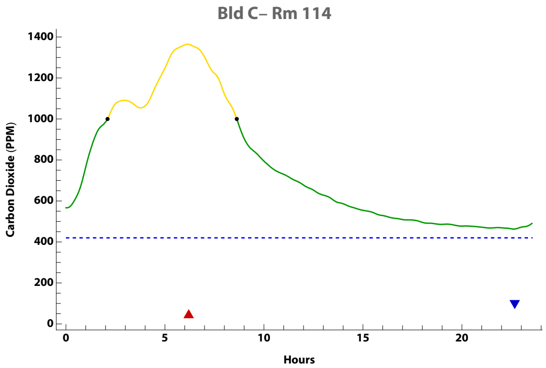


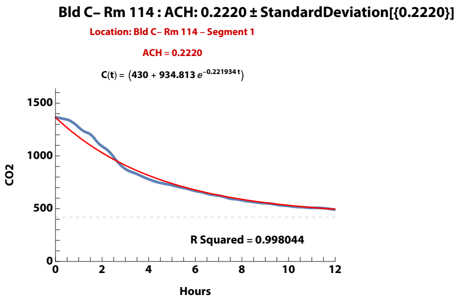


**Zand, et al. (2023). Ventilation during COVID–19 in a school for students with intellectual and developmental disabilities (IDD) [Submitted]**

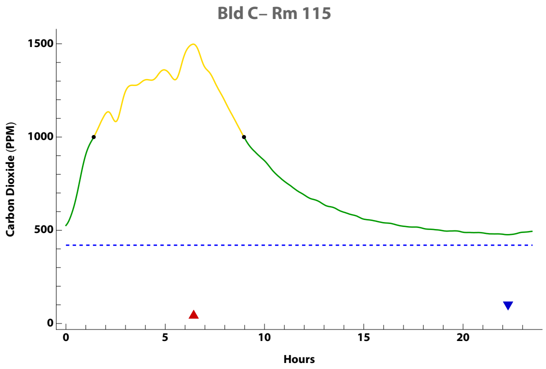


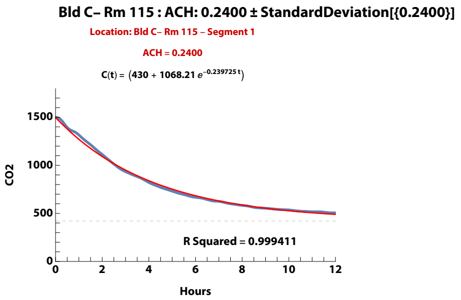


**Zand, et al. (2023). Ventilation during COVID–19 in a school for students with intellectual and developmental disabilities (IDD) [Submitted]**

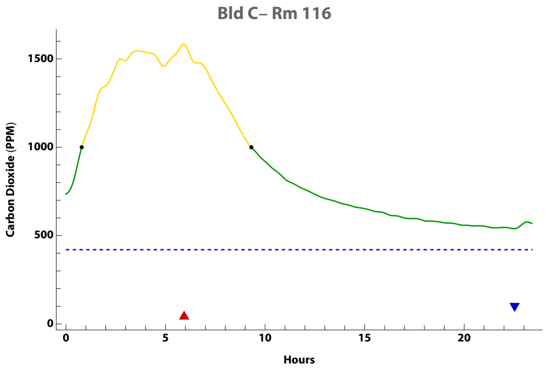


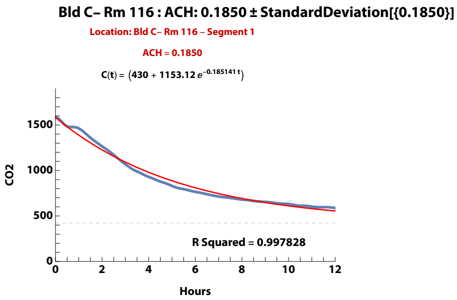


**Zand, et al. (2023). Ventilation during COVID–19 in a school for students with intellectual and developmental disabilities (IDD) [Submitted]**

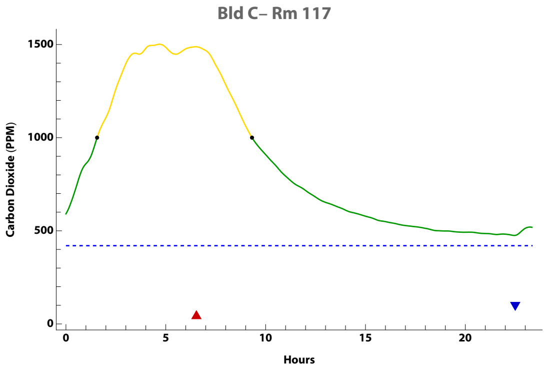


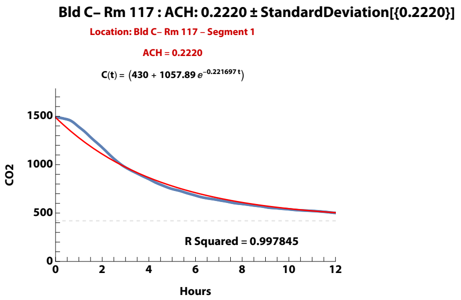


**Zand, et al. (2023). Ventilation during COVID–19 in a school for students with intellectual and developmental disabilities (IDD) [Submitted]**

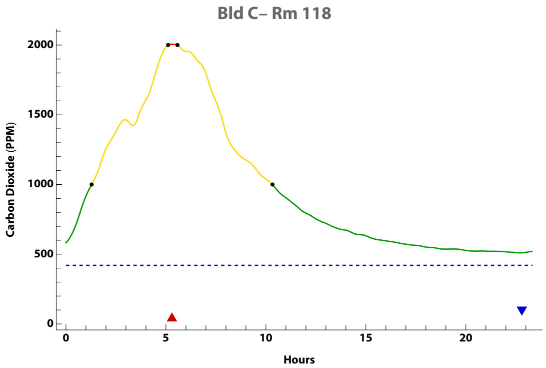


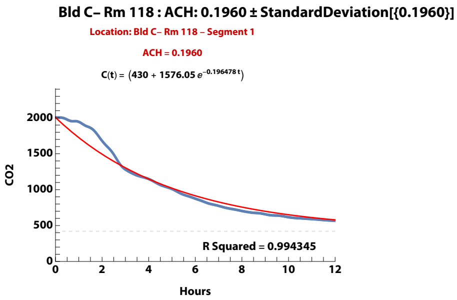


**Zand, et al. (2023). Ventilation during COVID–19 in a school for students with intellectual and developmental disabilities (IDD) [Submitted]**

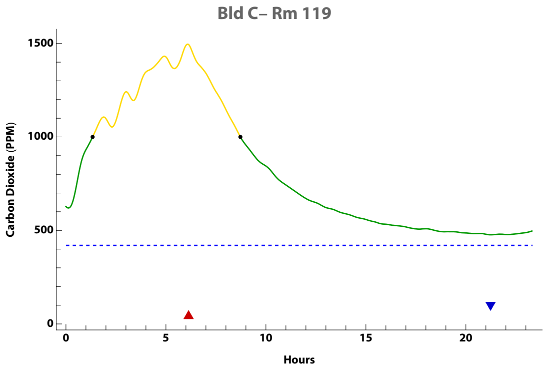


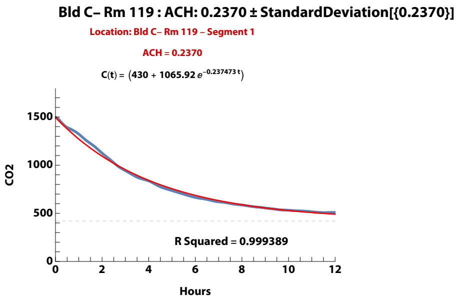


**Zand, et al. (2023). Ventilation during COVID–19 in a school for students with intellectual and developmental disabilities (IDD) [Submitted]**

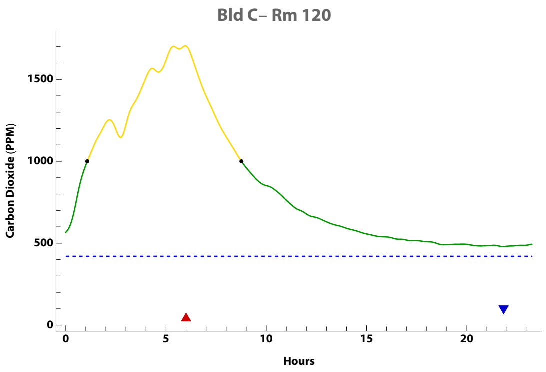


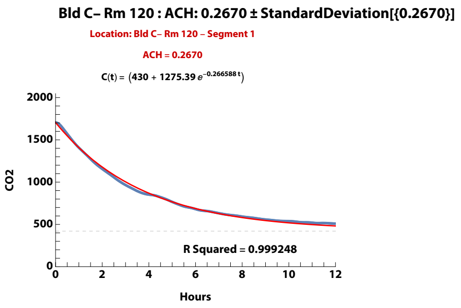


**Zand, et al. (2023). Ventilation during COVID–19 in a school for students with intellectual and developmental disabilities (IDD) [Submitted]**

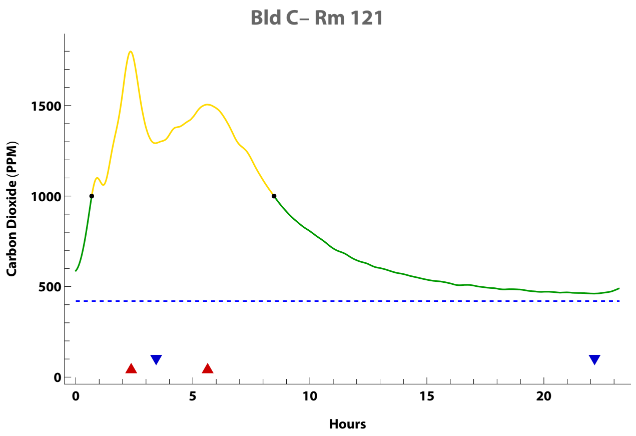


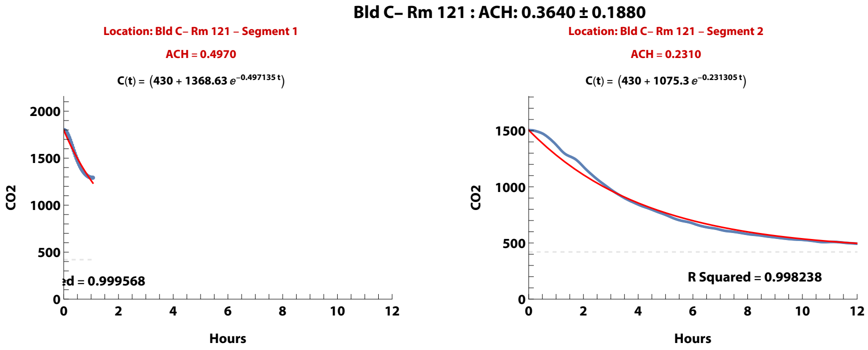


**Zand, et al. (2023). Ventilation during COVID–19 in a school for students with intellectual and developmental disabilities (IDD) [Submitted]**

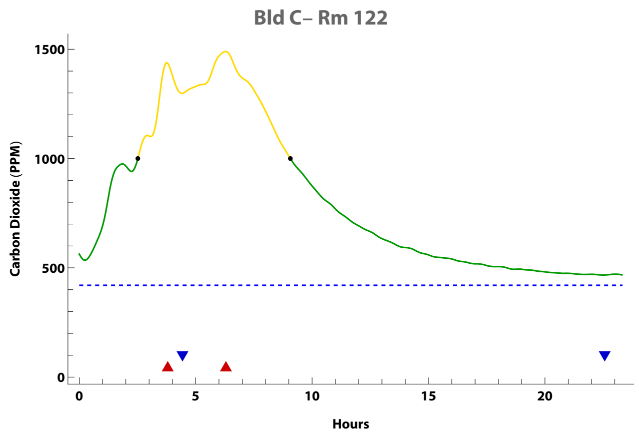


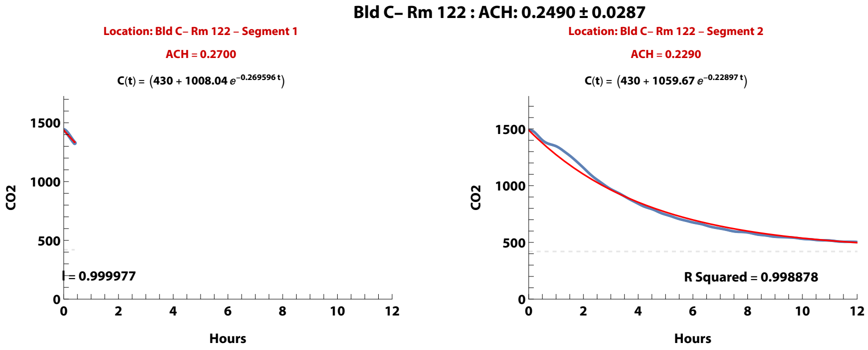


**Zand, et al. (2023). Ventilation during COVID–19 in a school for students with intellectual and developmental disabilities (IDD) [Submitted]**

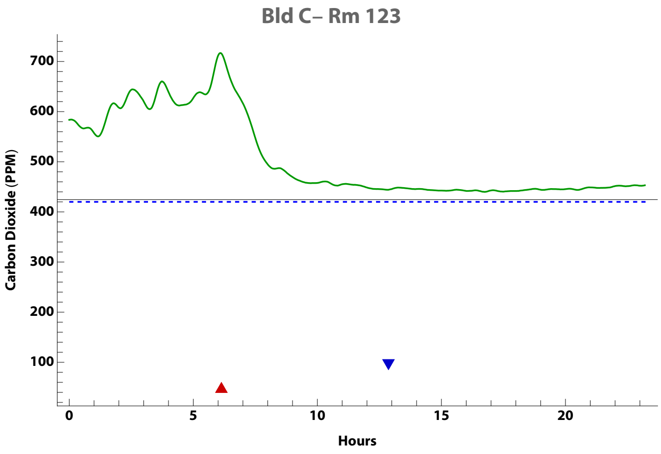


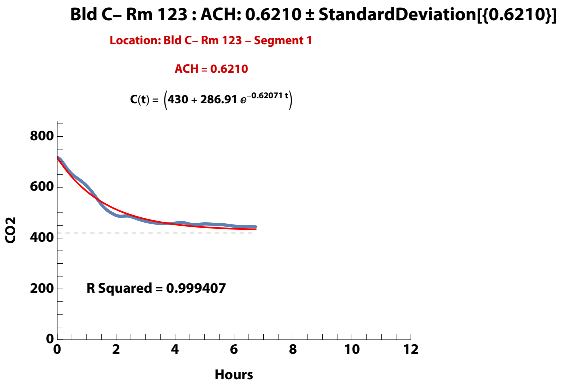


**Zand, et al. (2023). Ventilation during COVID–19 in a school for students with intellectual and developmental disabilities (IDD) [Submitted]**

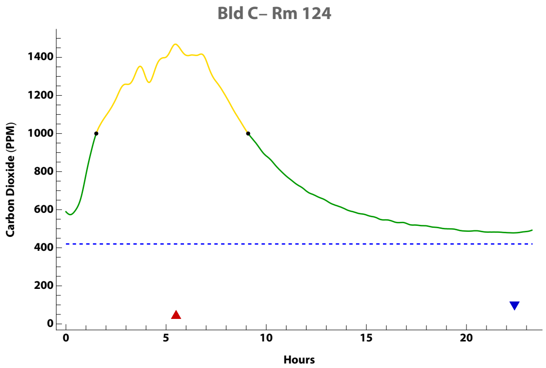


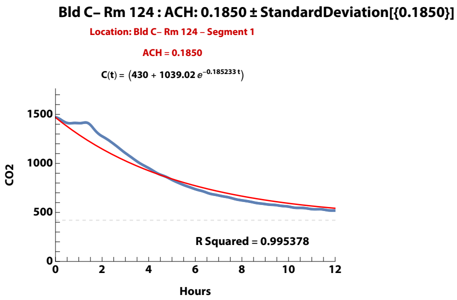


**Zand, et al. (2023). Ventilation during COVID–19 in a school for students with intellectual and developmental disabilities (IDD) [Submitted]**

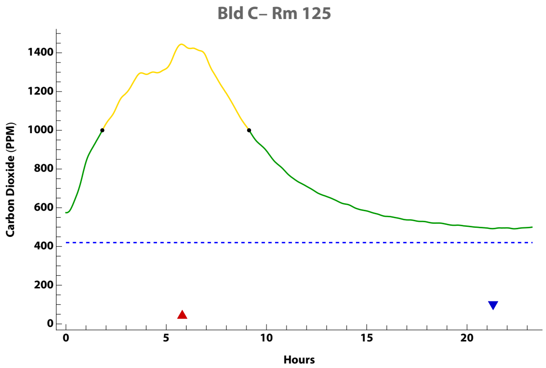


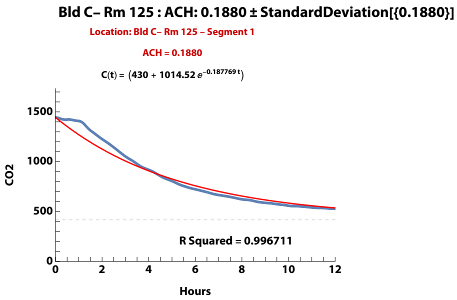


**Zand, et al. (2023). Ventilation during COVID–19 in a school for students with intellectual and developmental disabilities (IDD) [Submitted]**

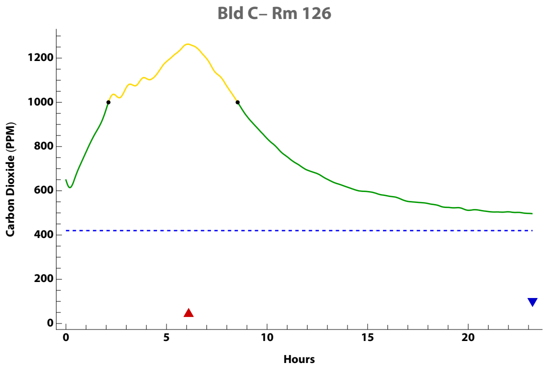


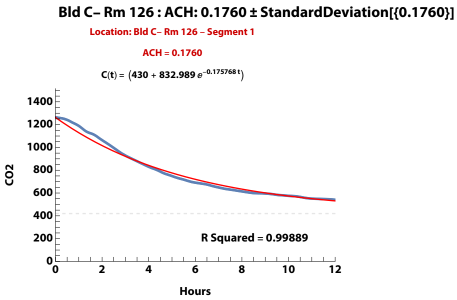


**Zand, et al. (2023). Ventilation during COVID–19 in a school for students with intellectual and developmental disabilities (IDD) [Submitted]**

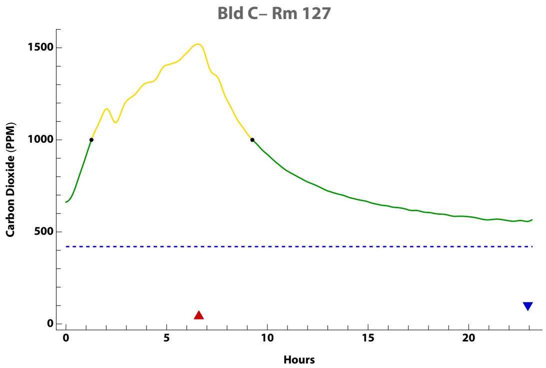


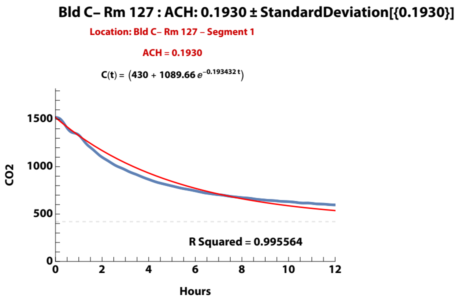


**Zand, et al. (2023). Ventilation during COVID–19 in a school for students with intellectual and developmental disabilities (IDD) [Submitted]**

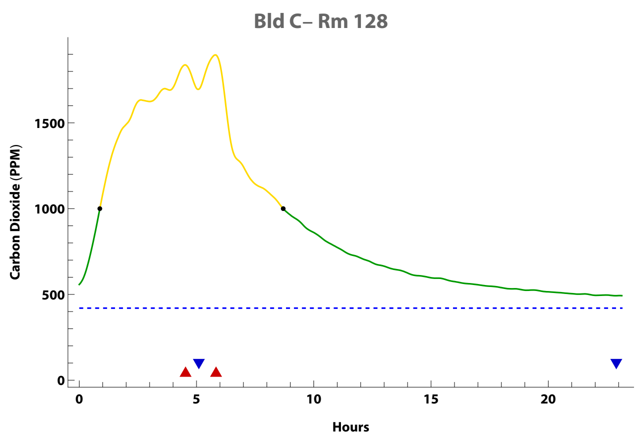


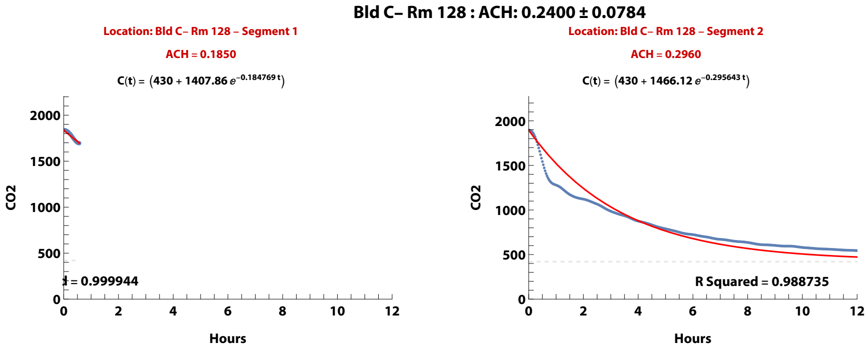


**Zand, et al. (2023). Ventilation during COVID–19 in a school for students with intellectual and developmental disabilities (IDD) [Submitted]**

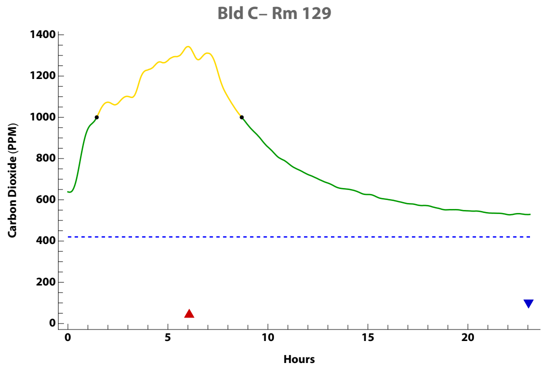


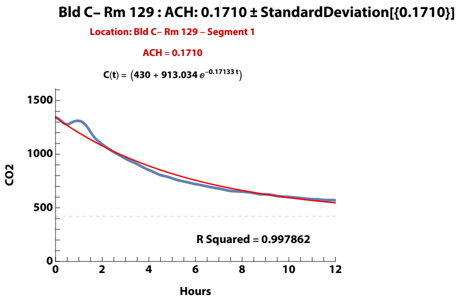


**Zand, et al. (2023). Ventilation during COVID–19 in a school for students with intellectual and developmental disabilities (IDD) [Submitted]**

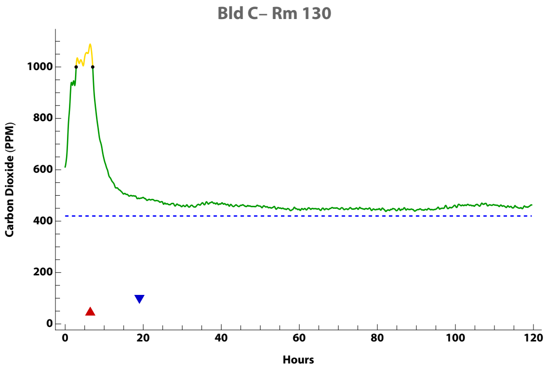


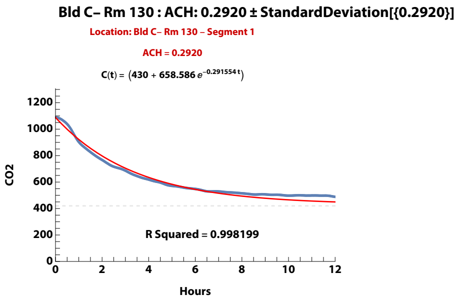


**Zand, et al. (2023). Ventilation during COVID–19 in a school for students with intellectual and developmental disabilities (IDD) [Submitted]**

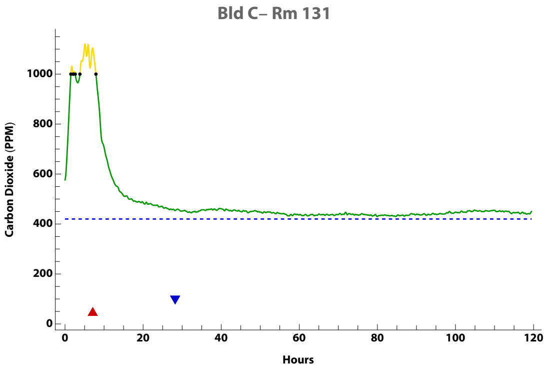


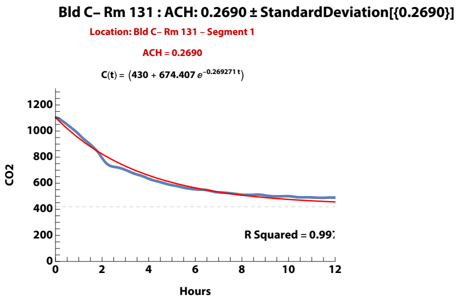


Zand, et al. (2023). Ventilation during COVID–19 in a school for students with intellectual and developmental disabilities (IDD) [Submitted]

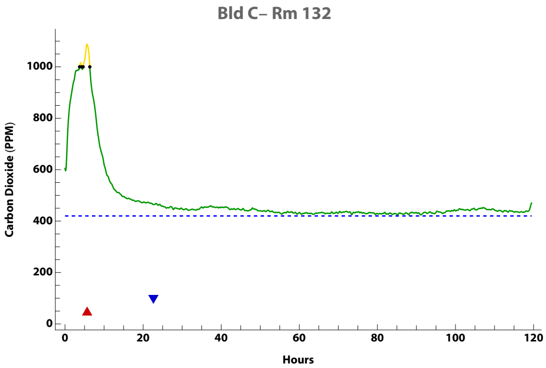


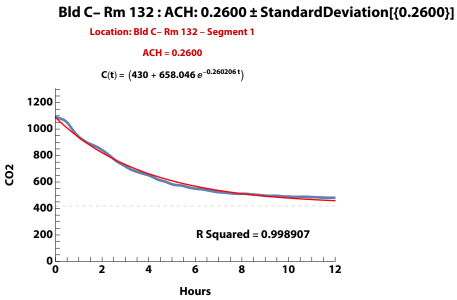


**Zand, et al. (2023). Ventilation during COVID–19 in a school for students with intellectual and developmental disabilities (IDD) [Submitted]**

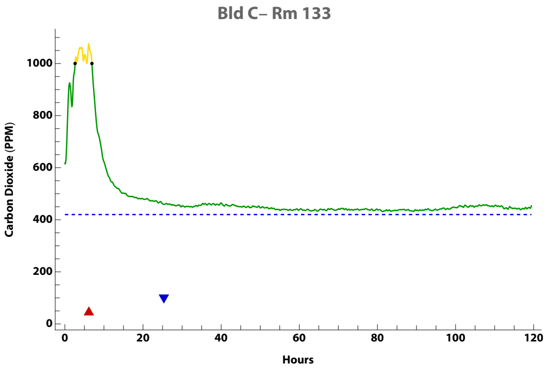


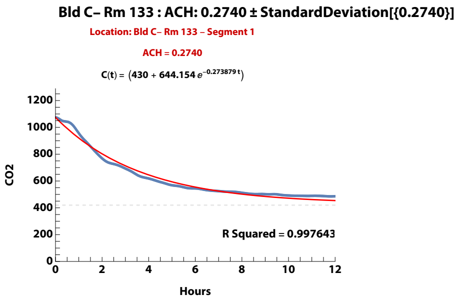


**Zand, et al. (2023). Ventilation during COVID–19 in a school for students with intellectual and developmental disabilities (IDD) [Submitted]**

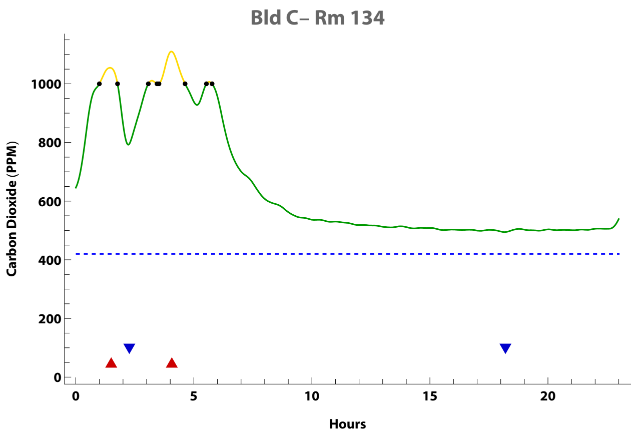


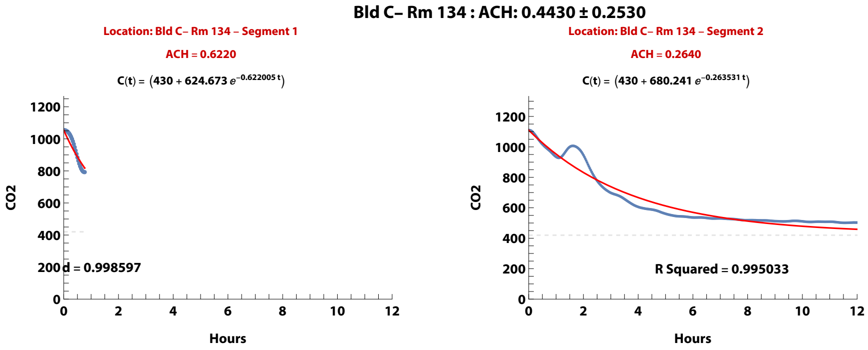


**Zand, et al. (2023). Ventilation during COVID–19 in a school for students with intellectual and developmental disabilities (IDD) [Submitted]**

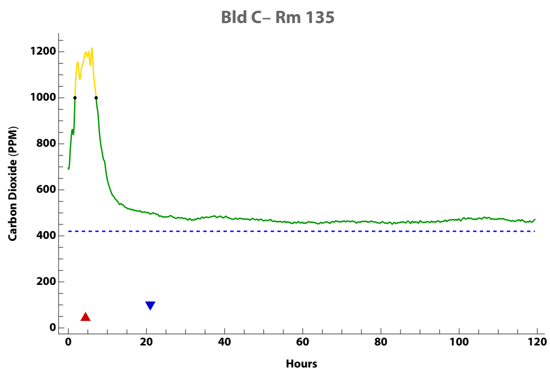


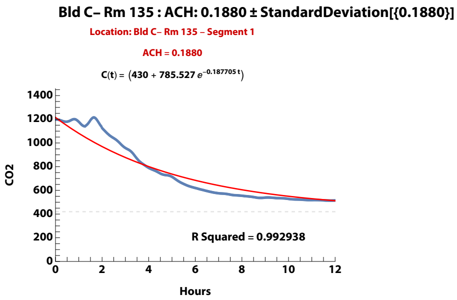


**Zand, et al. (2023). Ventilation during COVID–19 in a school for students with intellectual and developmental disabilities (IDD) [Submitted]**

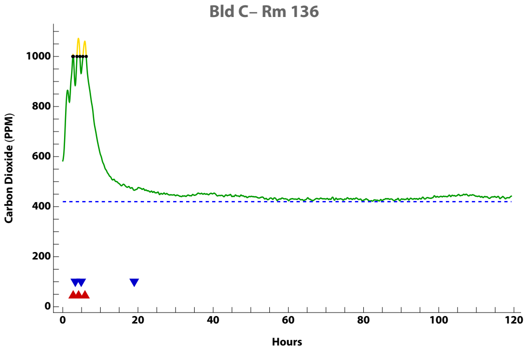


Bld C– Rm 136 : ACH: 0.3470 ± 0.0729

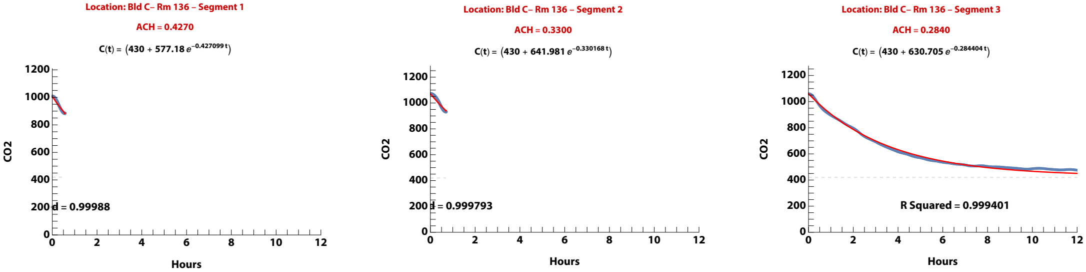


**Zand, et al. (2023). Ventilation during COVID–19 in a school for students with intellectual and developmental disabilities (IDD) [Submitted]**

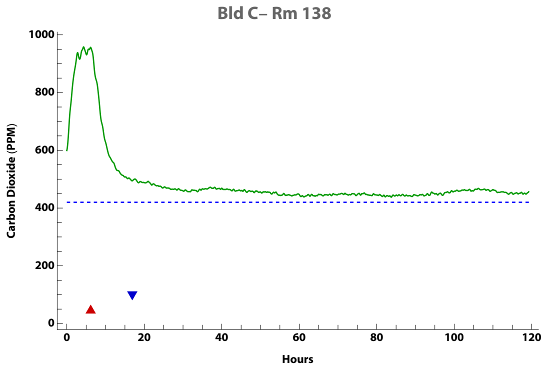


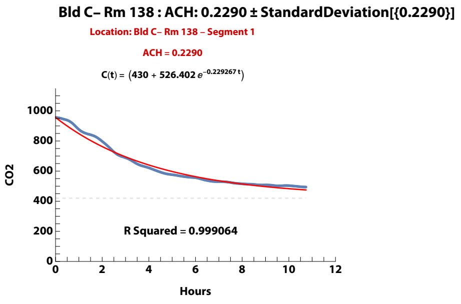


**Zand, et al. (2023). Ventilation during COVID–19 in a school for students with intellectual and developmental disabilities (IDD) [Submitted]**

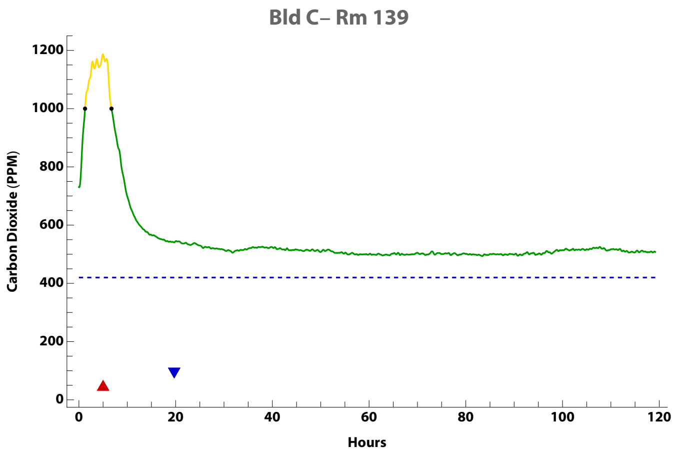


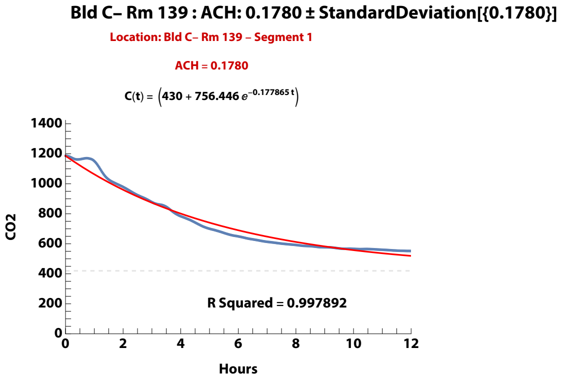


**Zand, et al. (2023). Ventilation during COVID–19 in a school for students with intellectual and developmental disabilities (IDD) [Submitted]**

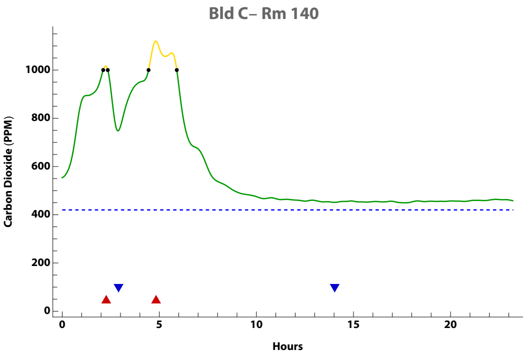


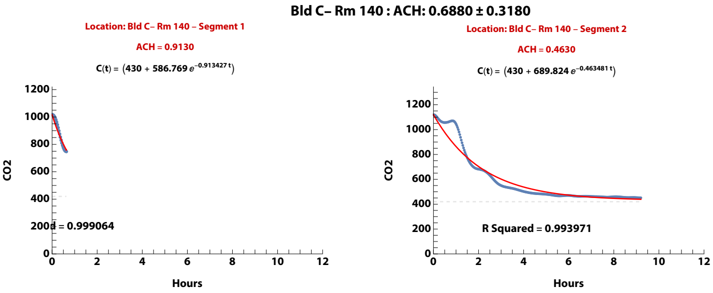


**Zand, et al. (2023). Ventilation during COVID–19 in a school for students with intellectual and developmental disabilities (IDD) [Submitted]**

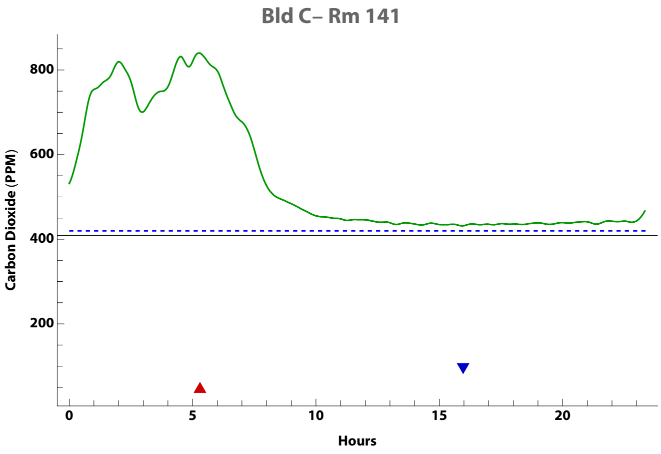


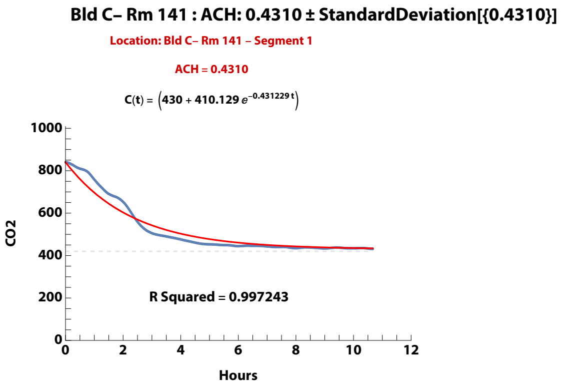


**Zand, et al. (2023). Ventilation during COVID–19 in a school for students with intellectual and developmental disabilities (IDD) [Submitted]**

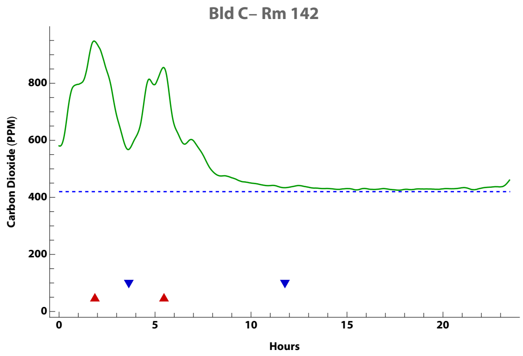


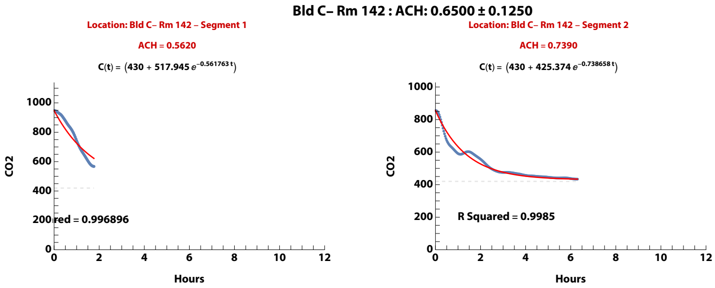


**Zand, et al. (2023). Ventilation during COVID–19 in a school for students with intellectual and developmental disabilities (IDD) [Submitted]**

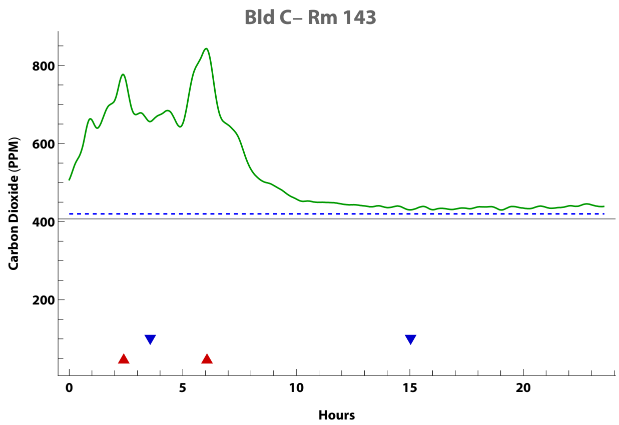


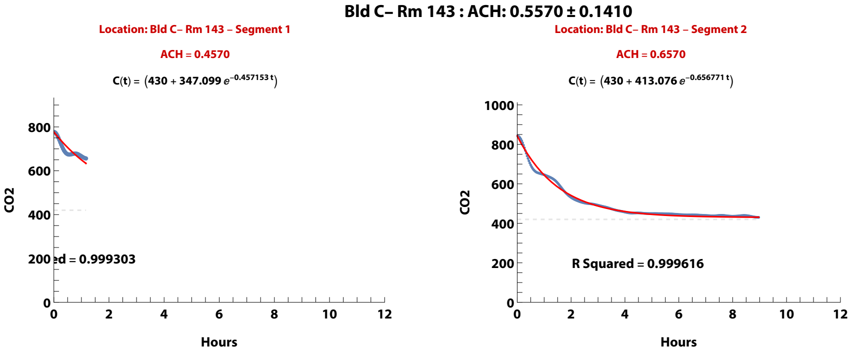


**Zand, et al. (2023). Ventilation during COVID–19 in a school for students with intellectual and developmental disabilities (IDD) [Submitted]**

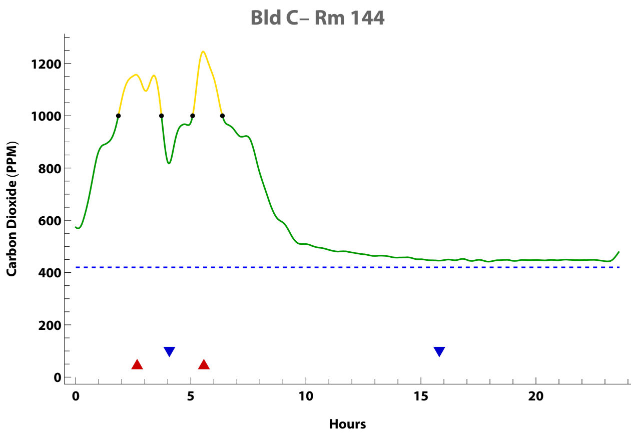


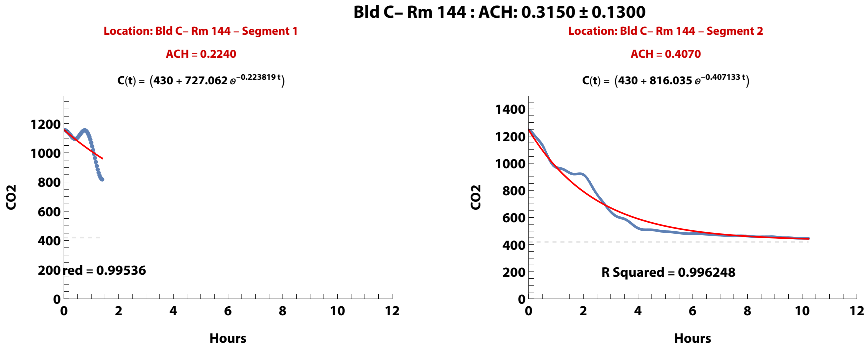


**Zand, et al. (2023). Ventilation during COVID–19 in a school for students with intellectual and developmental disabilities (IDD) [Submitted]**

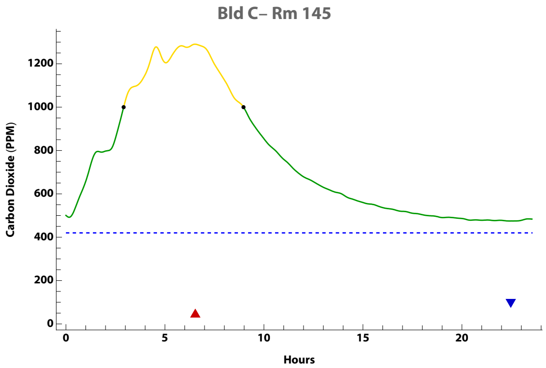


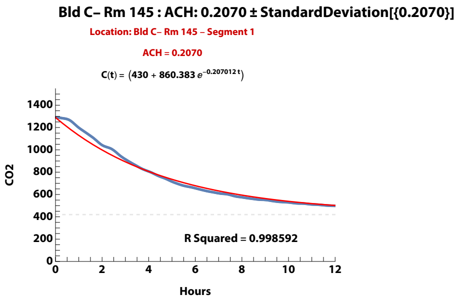


**Zand, et al. (2023). Ventilation during COVID–19 in a school for students with intellectual and developmental disabilities (IDD) [Submitted]**

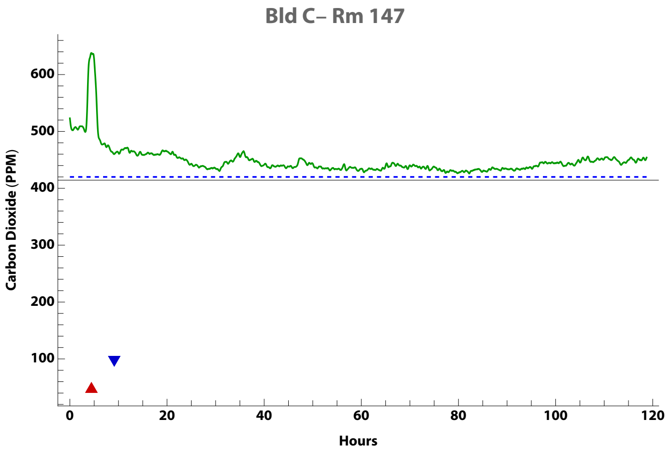


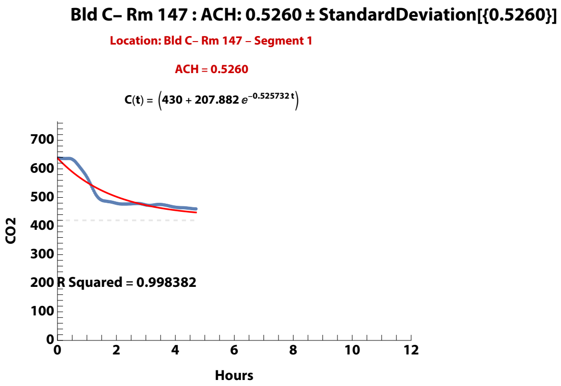


**Zand, et al. (2023). Ventilation during COVID–19 in a school for students with intellectual and developmental disabilities (IDD) [Submitted]**

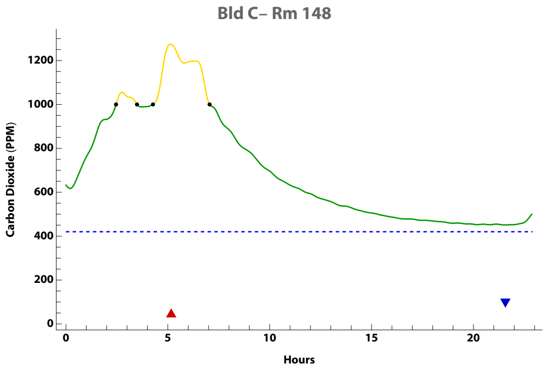


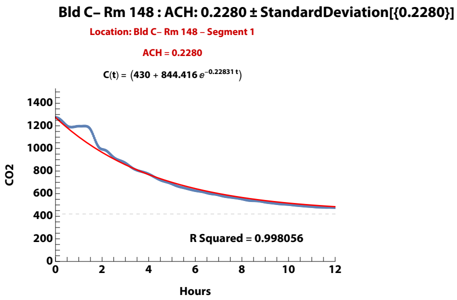


**Zand, et al. (2023). Ventilation during COVID–19 in a school for students with intellectual and developmental disabilities (IDD) [Submitted]**

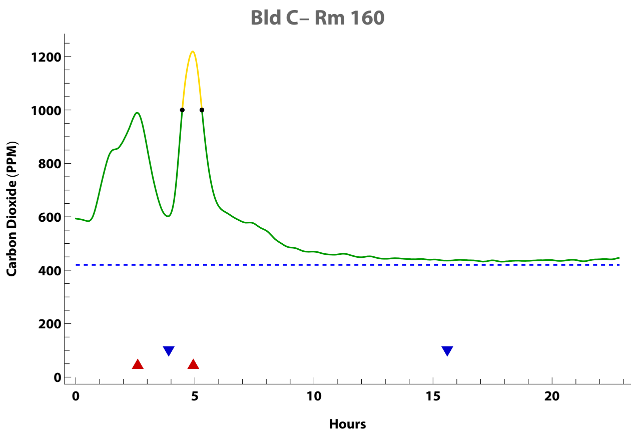


Bld C– Rm 160 : ACH: 0.8740 ± 0.0160

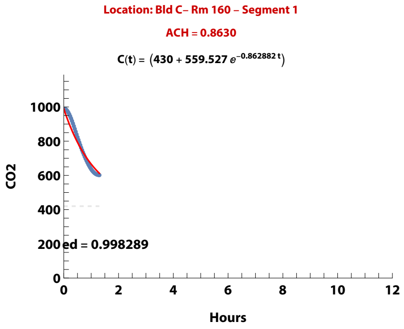


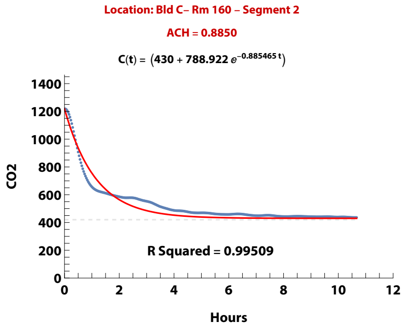


**Zand, et al. (2023). Ventilation during COVID–19 in a school for students with intellectual and developmental disabilities (IDD) [Submitted]**

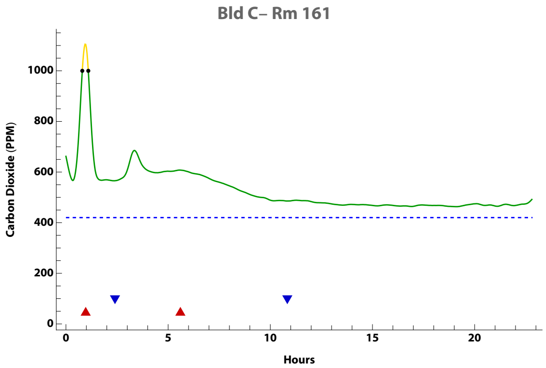


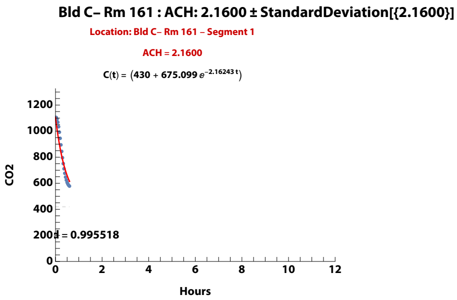


**Zand, et al. (2023). Ventilation during COVID–19 in a school for students with intellectual and developmental disabilities (IDD) [Submitted]**

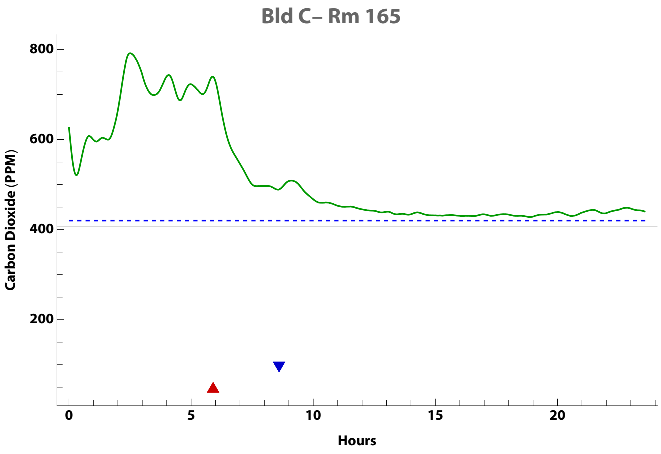


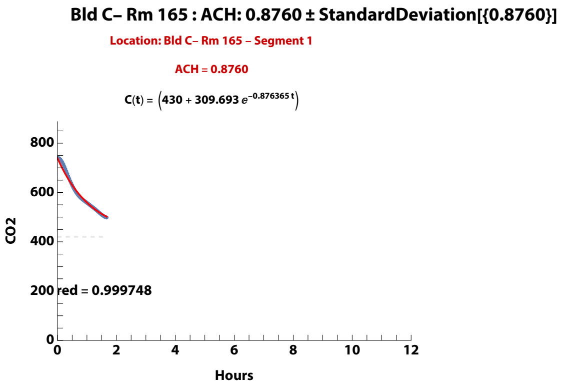


**Zand, et al. (2023). Ventilation during COVID–19 in a school for students with intellectual and developmental disabilities (IDD) [Submitted]**

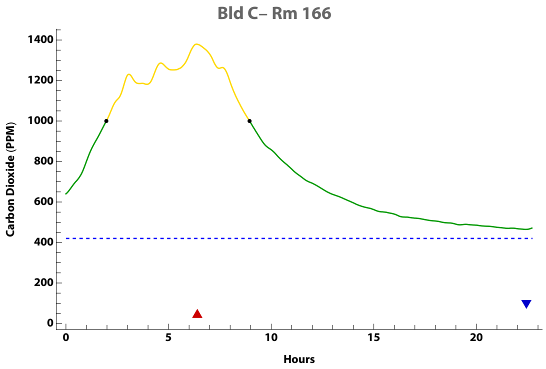


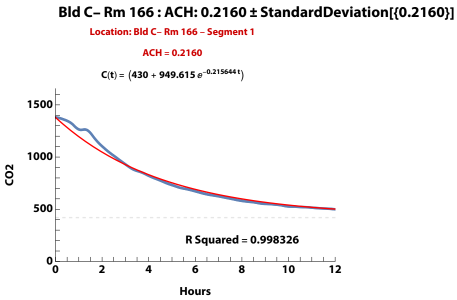


**Zand, et al. (2023). Ventilation during COVID–19 in a school for students with intellectual and developmental disabilities (IDD) [Submitted]**

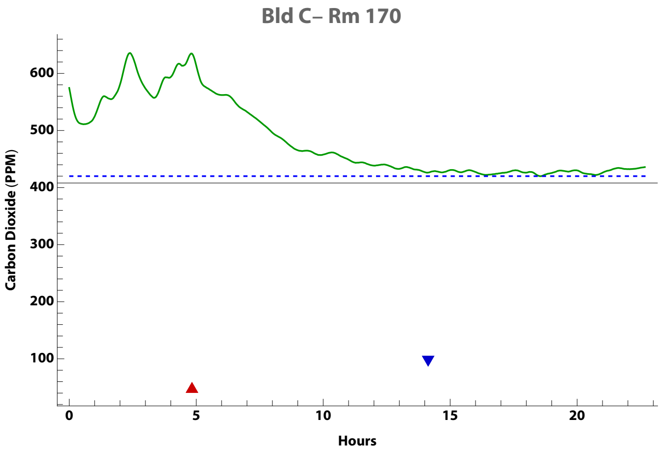


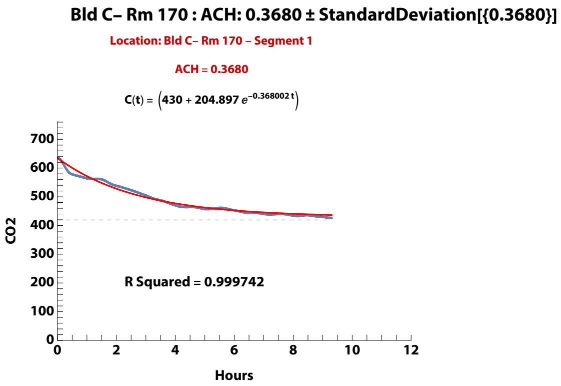


**Zand, et al. (2023). Ventilation during COVID–19 in a school for students with intellectual and developmental disabilities (IDD) [Submitted]**

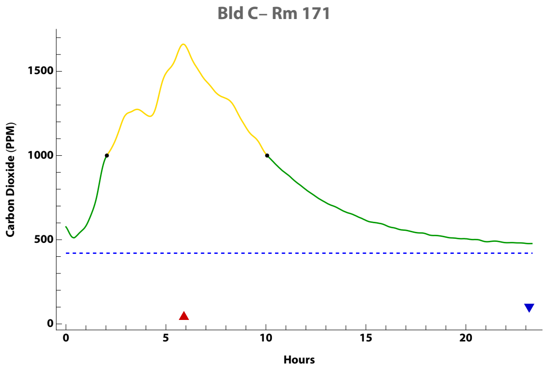


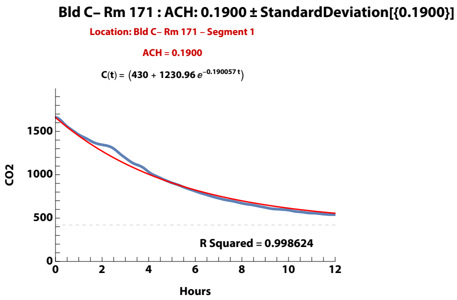


**Zand, et al. (2023). Ventilation during COVID–19 in a school for students with intellectual and developmental disabilities (IDD) [Submitted]**

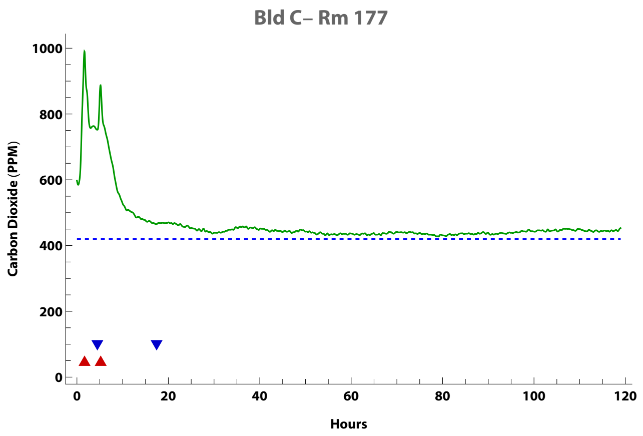


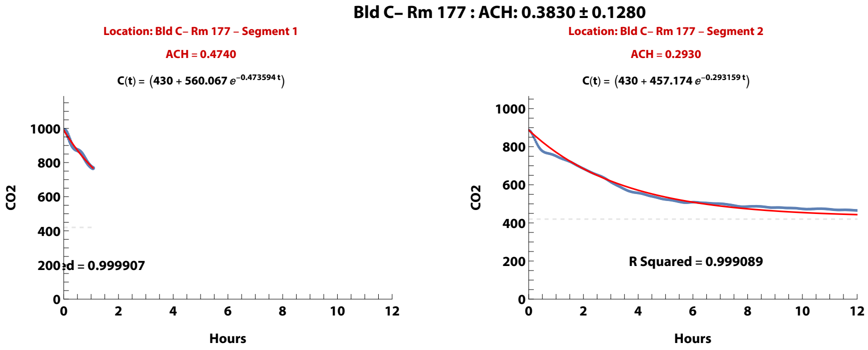


**Zand, et al. (2023). Ventilation during COVID–19 in a school for students with intellectual and developmental disabilities (IDD) [Submitted]**

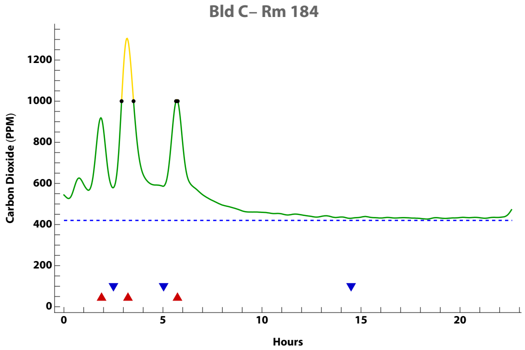


Bld C– Rm 184 : ACH: 1.6100 ± 0.2790

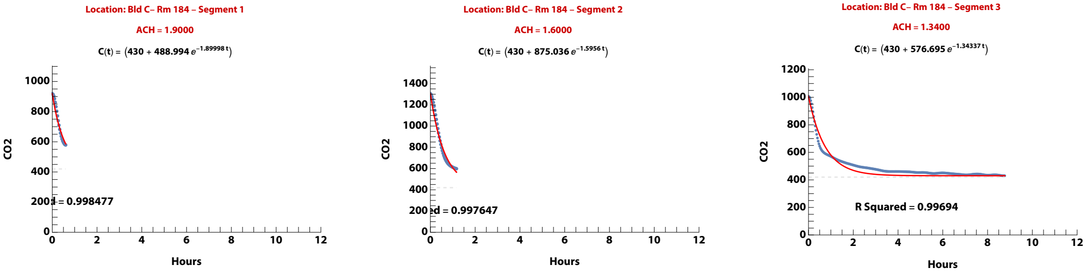


**Zand, et al. (2023). Ventilation during COVID–19 in a school for students with intellectual and developmental disabilities (IDD) [Submitted]**

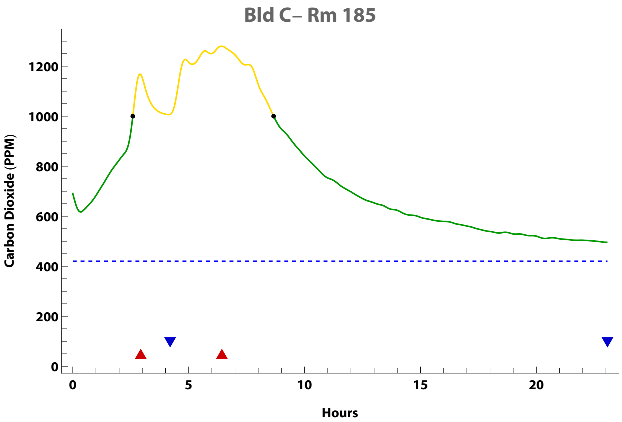


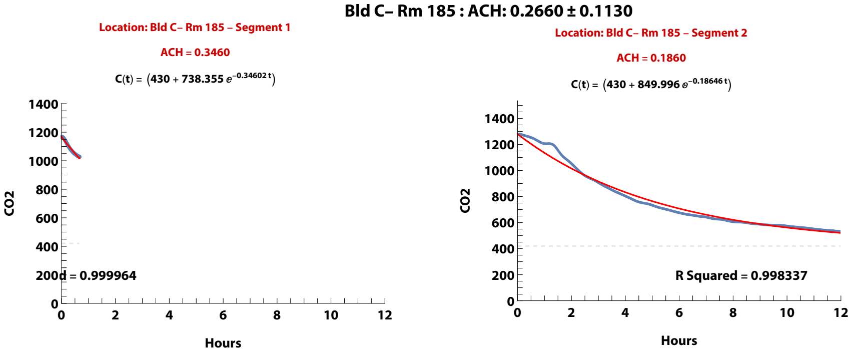


**Zand, et al. (2023). Ventilation during COVID–19 in a school for students with intellectual and developmental disabilities (IDD) [Submitted]**

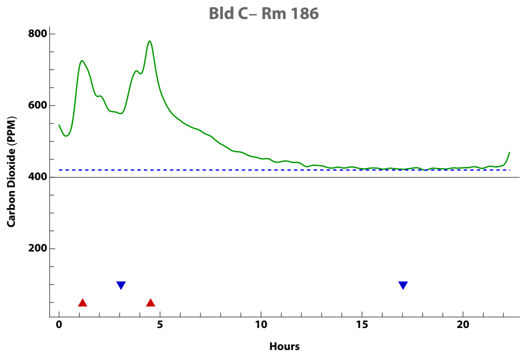


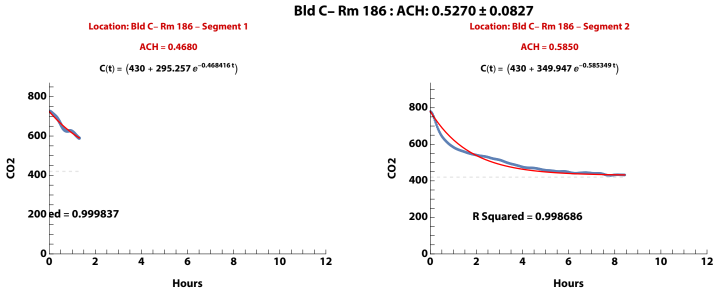


**Zand, et al. (2023). Ventilation during COVID–19 in a school for students with intellectual and developmental disabilities (IDD) [Submitted]**

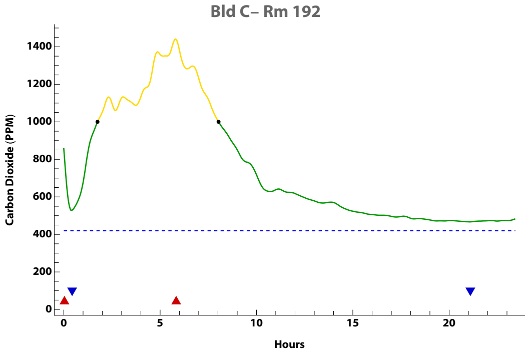


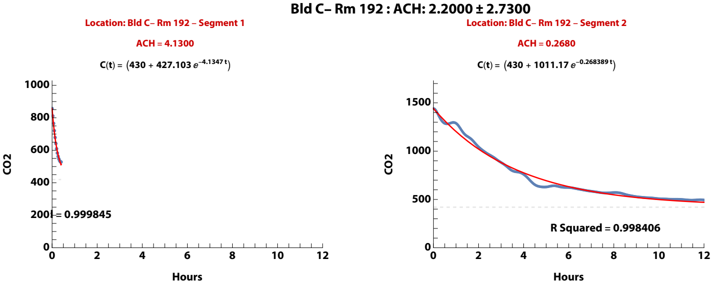


**Zand, et al. (2023). Ventilation during COVID–19 in a school for students with intellectual and developmental disabilities (IDD) [Submitted]**

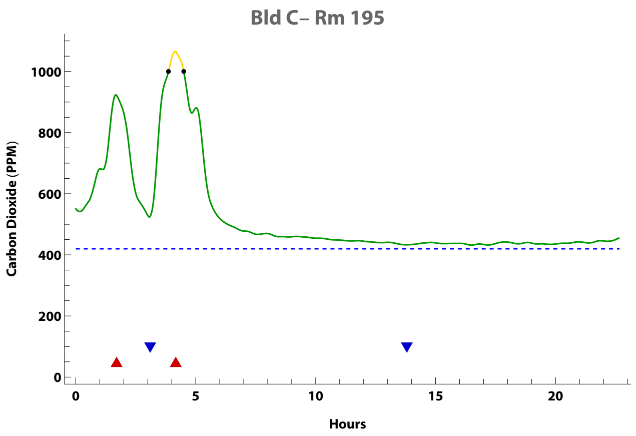


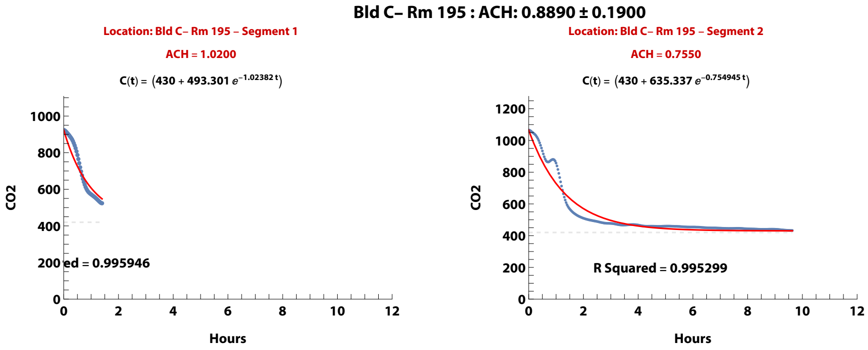


**Zand, et al. (2023). Ventilation during COVID–19 in a school for students with intellectual and developmental disabilities (IDD) [Submitted]**

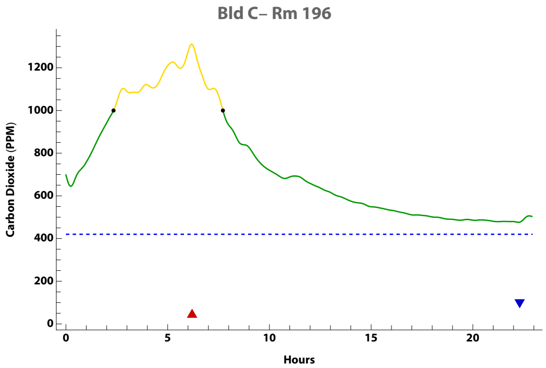


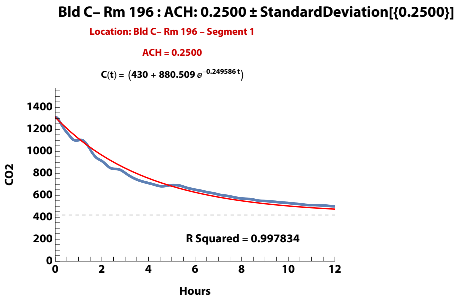


**Zand, et al. (2023). Ventilation during COVID–19 in a school for students with intellectual and developmental disabilities (IDD) [Submitted]**

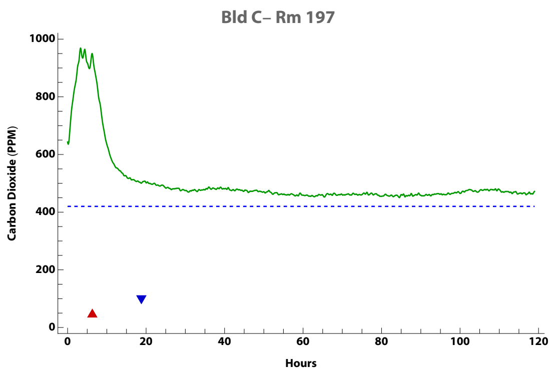


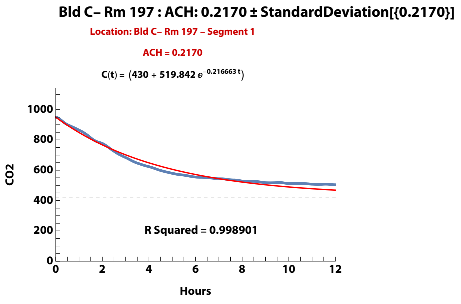


**Zand, et al. (2023). Ventilation during COVID–19 in a school for students with intellectual and developmental disabilities (IDD) [Submitted]**

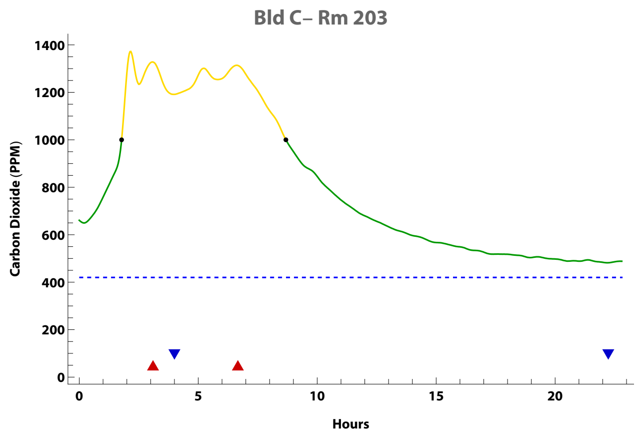


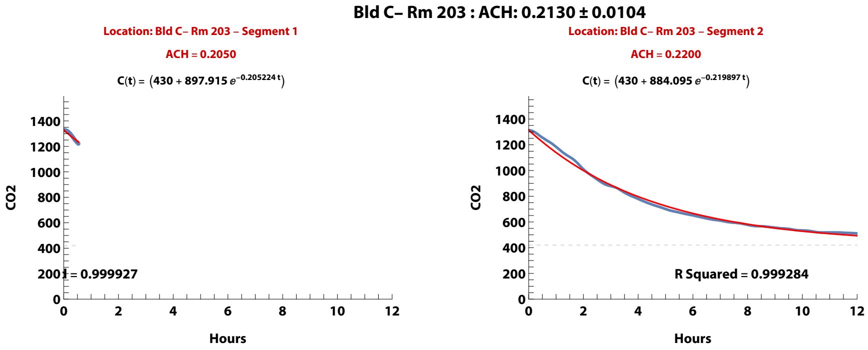


**Zand, et al. (2023). Ventilation during COVID–19 in a school for students with intellectual and developmental disabilities (IDD) [Submitted]**

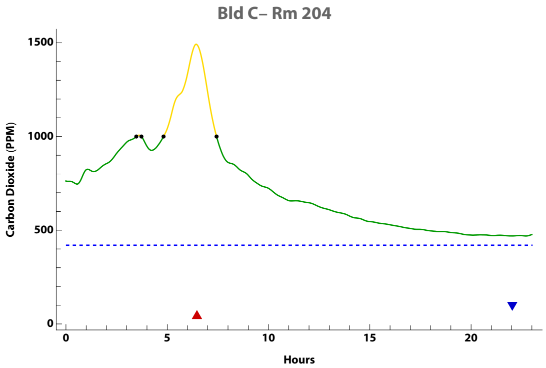


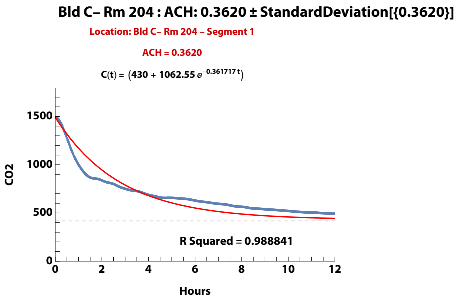


**Zand, et al. (2023). Ventilation during COVID–19 in a school for students with intellectual and developmental disabilities (IDD) [Submitted]**

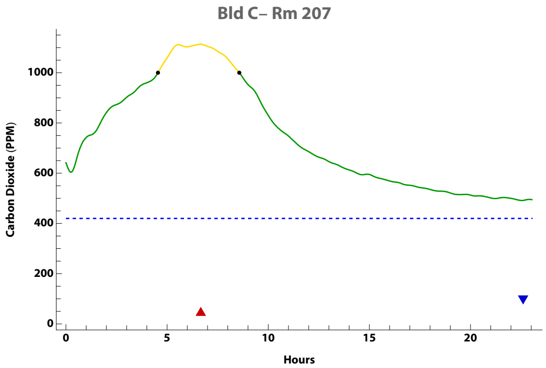


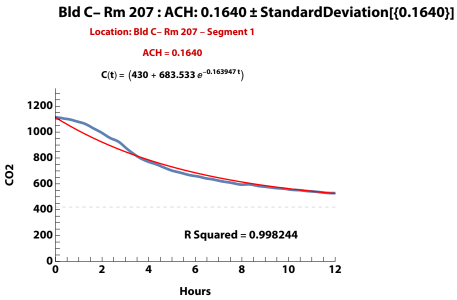


**Zand, et al. (2023). Ventilation during COVID–19 in a school for students with intellectual and developmental disabilities (IDD) [Submitted]**

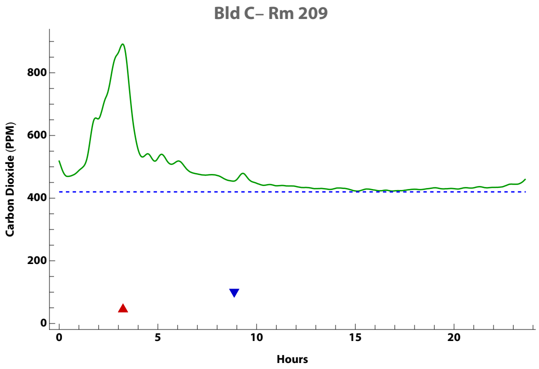


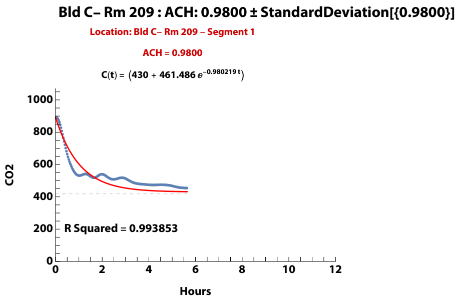


**Zand, et al. (2023). Ventilation during COVID–19 in a school for students with intellectual and developmental disabilities (IDD) [Submitted]**

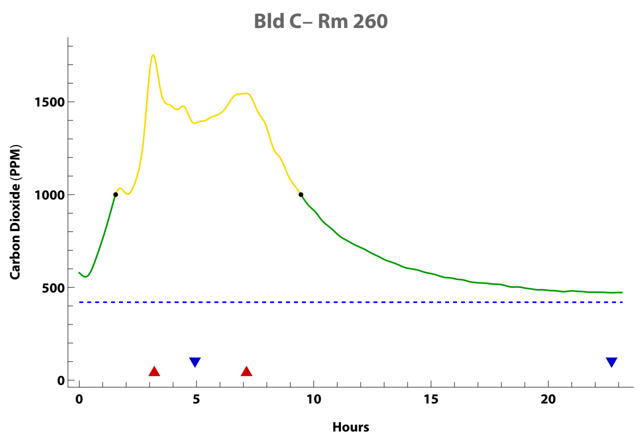


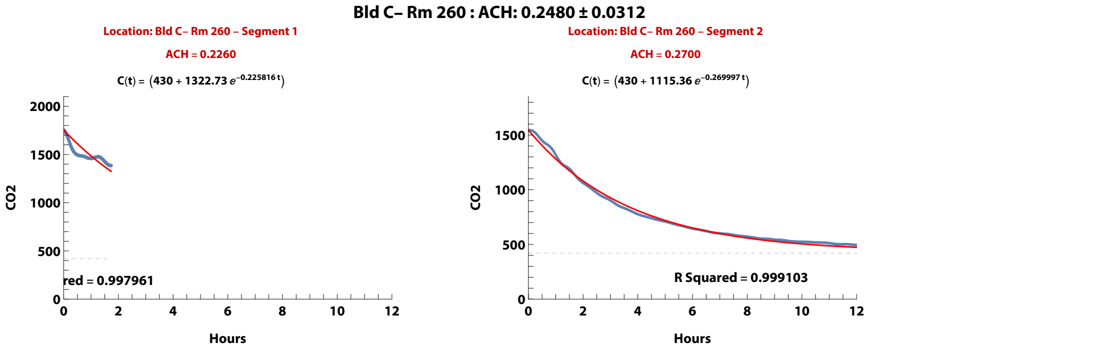


**Zand, et al. (2023). Ventilation during COVID–19 in a school for students with intellectual and developmental disabilities (IDD) [Submitted]**
